# Bodily self disturbances: a new clinical marker of persistent postural-perceptual dizziness

**DOI:** 10.64898/2026.01.30.26345201

**Authors:** Maélis Gobinet, Maya Elzière, Jacques Léonard, Christophe Lopez

**Author notes:** Correspondence to: Dr Christophe Lopez, Centre for Research in Psychology and Neuroscience (CRPN) – UMR 7077, Aix Marseille Univ & Centre National de la Recherche Scientifique (CNRS), Centre Saint-Charles, Fédération de Recherche 3C – Case B, 3, Place Victor Hugo. 13331 Marseille Cedex 03, France.

## Abstract

Persistent Postural-Perceptual Dizziness (PPPD) is among the most prevalent chronic neuro-otologic disorders, affecting 15–20% of adults seen in neurology and specialized dizziness clinics. Classified as a functional vestibular disorder and defined by established diagnostic criteria, PPPD typically follows peripheral or central otoneurological disorders. However, the mechanisms underlying the transition from these disorders to chronic perceptual dizziness remain unclear. Beyond maladaptive postural control and visual dependence, theoretical models implicate altered multisensory integration and disrupted predictive processing. Such mechanisms may extend to disturbances of the bodily self, a dimension increasingly recognized in functional neurological disorders, but not yet systematically investigated in PPPD.

We characterized bodily self disturbances in PPPD by assessing depersonalization-derealization symptoms in a large cross-sectional study (*n* = 455), including 100 patients with PPPD, 180 patients with other otoneurological disorders, and 175 healthy controls. Depersonalization-derealization symptoms were assessed using the Cambridge Depersonalization Scale, alongside measures of anxiety, depression, dizziness-related impairment, and PPPD symptom severity. PPPD patients exhibited markedly elevated depersonalization-derealization symptoms compared to both other otoneurological disorders and healthy controls (all *P* < 0.001). Notably, 20% of PPPD patients met the threshold for clinical depersonalization-derealization, compared with 7.2% of other otoneurological disorders and <1% of controls. Depersonalization-derealization severity in PPPD overlapped with levels observed in other functional neurological disorders but remained lower than in primary dissociative disorders.

Factor analyses identified three depersonalization-derealization dimensions: *Bodily Self Disturbances*, *Cognitive and Affective Detachment*, and *Numbing*. Only *Bodily Self Disturbances,* capturing disruptions in self-location, agency, body ownership, and first-person perspective, robustly differentiated PPPD from other otoneurological disorders (*η*² = 0.20, *P* < 0.001). This dimension predicted PPPD diagnosis (odds ratio = 1.40, *P* < 0.001), and showed significant discriminative ability (AUC = 0.66). Individuals in the highest decile of *Bodily Self Disturbances* had nearly tenfold increased odds of PPPD. Structural equation modelling confirmed a direct effect of PPPD on *Bodily Self Disturbances*, partially mediated by depressive symptoms but independent of age, sex, migraine, and anxiety.

These findings identify depersonalization-derealization as a previously unrecognized component of the PPPD phenotype and establish bodily self disturbances as a novel clinical marker for PPPD, refining phenotyping and informing pathophysiological models.

## Introduction

Persistent Postural-Perceptual Dizziness (PPPD) is now among the most prevalent chronic neuro-otologic conditions, representing approximately 15−20% of adult seen in neurology and specialized dizziness clinics.^1,2^ The Bárány Society^3^ and ICD-11^4^ define PPPD by persistent non-vertiginous dizziness and/or unsteadiness, present on most days for at least three months. Symptoms are exacerbated by upright posture, active or passive movement, and exposure to complex or moving visual patterns.^1,3^

Conceptualized as a *functional vestibular disorder*, PPPD integrates earlier diagnostic entities including phobic postural vertigo, visual vertigo, space-motion discomfort, and chronic subjective dizziness.^3,5,6^ PPPD is most often precipitated by peripheral or central otoneurological disorders (OD) such as benign paroxysmal positional vertigo, vestibular migraine, or acute unilateral vestibulopathy.^3,7,8^

Why only some patients with vestibular disorders develop PPPD remains unclear. Current pathophysiological models^1,6,9,10^ implicate maladaptive postural control, excessive visual dependence, motion hypersensitivity and psychological factors as key contributors.^1,10^ Consistent with this, PPPD patients generally report higher levels of anxiety and depression than those with other OD.^11,12^ Trait anxiety^1,7,12^, neuroticism^13,14^, and pre-existing symptoms^15^ may confer susceptibility to PPPD, whereas state anxiety and hypervigilance towards bodily sensations and balance may exacerbate symptoms^1,12^, perpetuating a maladaptive vicious cycle.^1,3,10^ Another suggested mechanism is multisensory integration dysfunction.^10,16–18^ Beyond these mechanisms, PPPD shares features with other functional neurological disorder^19^ and may be conceptualized within a predictive coding framework. This framework proposes that the brain continuously generates predictions about bodily states and compares them to incoming sensory inputs, constructing the sense of agency over bodily events during movement.^20,21^ Disturbances in this inferential process, whether due to imprecise predictions or mismatches between prediction and sensory feedback^19,22–24^, may disrupt experiences of the body and surroundings. This could lead not only to the symptoms observed in PPPD but also to alterations in the bodily self. The bodily self refers to a minimal, pre-reflective form of selfhood^25^ encompassing four core phenomenal dimensions: *self-location* (the sense of occupying a specific volume of space, normally within one’s physical boundaries), *body ownership* (the feeling that one’s own body belong to oneself), *first-person perspective* (the egocentric viewpoint from which the world is experienced), and *agency* (the sense of initiating and controlling one’s own actions).

Alterations in these dimensions have long been reported in OD^26–28^, described as depersonalization-derealization (DD), involving feelings of unreality, detachment from the self, and altered perception of the environment.^29,30^ DD symptoms are common across psychiatric, neurological, and functional disorders^31–34^, and are more prevalent in OD than in the general population.^35–38^ In OD, disturbances of the bodily self may manifest as out-of-body experiences^44^ or distortions of the body image^40^, including Alice in Wonderland syndrome.^41^ DD symptoms also show strong association with anxiety and depression.^42,43^

Despite these observations, it remains unknown whether PPPD patients experience more severe bodily self disturbances than patients with other OD. Recent semi-structured interviews in a small sample of patients identified themes of identity loss, dissociation, and depersonalization in PPPD^44^, suggesting that alterations of the bodily self may form part of its clinical phenotype. Establishing these disturbances could advance understanding of PPPD pathophysiology and provide useful clinical markers.

Here, we assessed depersonalization-derealization symptoms in a large sample of individuals with PPPD, other OD, and healthy controls. Our specific objectives were to: (i) characterize the profile of DD symptoms in PPPD; (ii) evaluate their prevalence and severity relative to other OD, as well as psychiatric and functional neurological disorders; (iii) assess their predictive value in differentiating PPPD from OD; and (iv) examine whether anxiety and depression mediate the association between PPPD and DD symptoms, while controlling for age, sex, and migraine.

## Materials and methods

This cross-sectional observational study adhered to the STROBE guidelines.^45^

### Participants

Participants were consecutively enrolled (March 2022–October 2024) in the PERSOCOR cohort, a monocentric prospective study conducted at the European Hospital (Marseille, France). Eligible patients were ≥18 years old, presented with vertigo/dizziness of any aetiology, and had no severe neurological or psychiatric comorbidities unrelated to the otoneurological disorder. Healthy controls (HC), recruited among patients’ relatives and hospital staff, reported no lifetime history of vertigo/dizziness, or neurological/psychiatric disorders. Study procedures were explained identically to all participants.

After excluding participants with multiple missing data and revised diagnosis, 455 individuals (100 PPPD, 180 OD, 175 HC) were retained for the final analysis following age-and sex-matching (**Fig. 1a)**. PPPD was diagnosed according to Bárány Society criteria.^3^ Clinical stratification was based on detailed otoneurological assessment and medical history.

**Figure 1.**
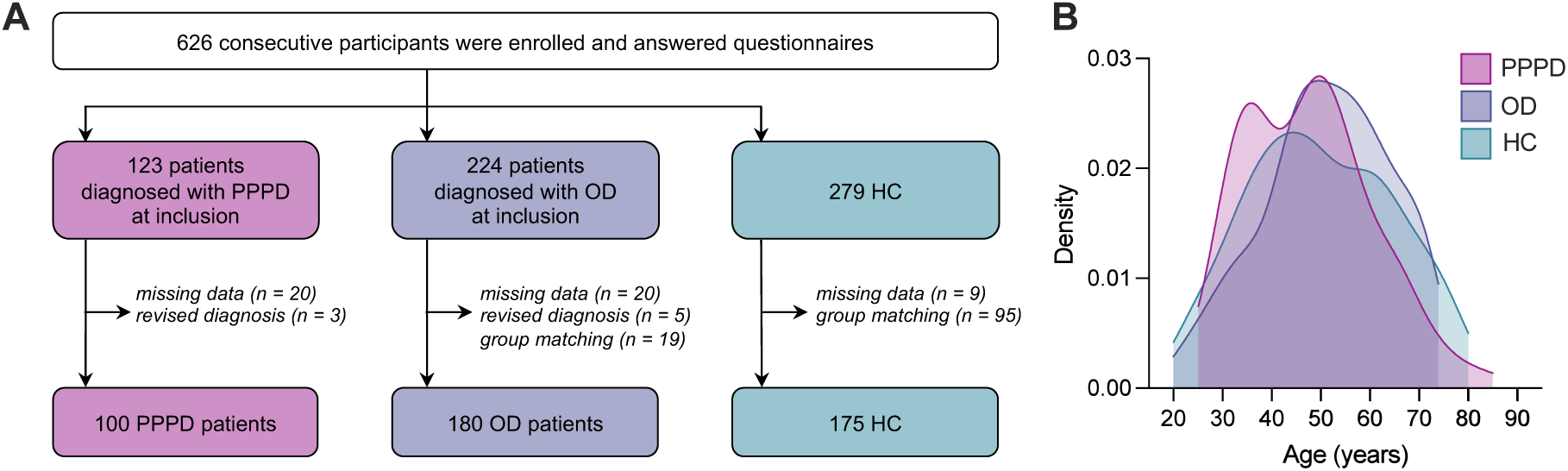
Participants inclusion and age distribution. **(A)** Flowchart of participants inclusion. **(B)** Kernel density plots showing similar age distribution in three participant groups. PPPD = persistent postural-perceptual dizziness; OD = otoneurological disorders; HC = healthy controls.

### Ethics approval

The study was approved by the Ethics Committee (CPP Île de France II, n°2021-A03111-40) and conducted in accordance with the Declaration of Helsinki. All participants provided written informed consent and received no compensation.

### Measures

#### Sociodemographic and clinical variables

Demographic data were collected, including age, sex, education, marital and employment status. We also recorded information about smoking and alcohol habits, migraine history, disease history and duration, as well as current antidepressant treatment (**Table 1**, **Supplementary Table 2**).

**Table 1.** Demographic and clinical characteristics. Highest education level was coded according to the French education system: Level 1 = before high school; Level 2 = high school diploma; Level 3 = two years after high school; Level 4 = Bachelor’s degree, Level 5 = Master’s degree, Engineering degree, PhD, or MD. * Statistically significant differences (*P <* 0.05) are shown in bold.

|  | PPPD<br>(*n* = 100) | OD<br>(*n* = 180) | HC<br>(*n* = 175) | Statistics |
| --- | --- | --- | --- | --- |
| Age, mean ± SD (years) | 47.6 ± 12.8 | 51.3 ± 12.8 | 49.7 ± 14.9 | $F(2) = 2.35, P = 0.10$ |
| Sex, *n* (%) | | | | $\chi^2(2) = 5.49, P = 0.064$ |
| *Females* | 75 (75%) | 120 (67%) | 107 (61%) |  |
| *Males* | 25 (25%) | 60 (33 %) | 68 (39%) |  |
| Handedness, *n* (%) | | | | $\chi^2(2) = 0.80, P = 0.93$ |
| *Right-handed* | 87 (92%) | 159 (90%) | 157 (92%) |  |
| *Left-handed* | 8 (8%) | 16 (9%) | 13 (8%) |  |
| *Ambidextrous* | 0 (0%) | 1 (1%) | 1 (1%) |  |
| Education level, % | | | | $\chi^2(8) = 10.34, P = 0.242$ |
| *Level 1* | 17.0% | 31.3% | 23.6% |  |
| *Level 2* | 17.0% | 12.9% | 19.0% |  |
| *Level 3* | 24.0% | 24.0% | 24.7% |  |
| *Level 4* | 19.0% | 15.0% | 13.2% |  |
| *Level 5* | 23.0% | 16.8% | 19.5% |  |
| Employment status, % | | | | $\chi^2(6) = 12.59, P = 0.050$ |
| *Employed* | 75.2% | 71.0% | 69.1% |  |
| *Student* | 1.1% | 0.6% | 4.6% |  |
| *Retired* | 12.9% | 20.7% | 21.7% |  |
| *Unemployed* | 10.8% | 7.7% | 4.6% |  |
| Marital status, % | | | | $\chi^2(4) = 10.15, P < 0.05$ |
| *Single* | 21.0% | 20.7% | 15.5% |  |
| *Married/couple* | 60.0% | 64.3% | 75.9% |  |
| *Divorced/widowed* | 19.00% | 15.0% | 8.6% |  |
| Smokers, % | 26.0% | 17.3% | 22.9% | $\chi^2(2) = 3.26, P = 0.196$ |
| Alcohol consumption, % | | | | $\chi^2(6) = 18.04 P < 0.01$ |
| *None* | 72.7% | 61.7% | 50.0% |  |
| *1 to 5 units/week* | 26.3% | 34.4% | 44.2% |  |
| *6 to 10 units/week* | 0.0% | 3.9% | 5.2% |  |
| *>10 units/week* | 1.0% | 0.0% | 0.6% |  |
| Migraine, % | 38.0% | 36.9% | 19.7% | $\chi^2(2) = 15.48, P < 0.001$ |

#### Otoneurological assessment

Patients underwent standard vestibular and auditory assessments according to their clinical presentation, including videonystagmography (spontaneous and positional nystagmus, head-shaking test [HST]; Synapsys), pendular rotatory test (Synapsys), caloric testing (Synapsys), video head impulse test (vHIT; Otometrics), cervical vestibular-evoked myogenic potentials (cVEMPs; Interacoustics), pure-tone audiometry (Interacoustics), and contrast-enhanced MRI. Clinical data acquired either at inclusion or from prior assessments were extracted from medical records. HC participants did not undergo otoneurological testing.

#### Depersonalization-derealization

DD symptoms were assessed with the 29-item Cambridge Depersonalization Scale (CDS).^46^ Participants rated the frequency (4-point scale) and duration (6-point scale) of each symptom over the past six months, giving a score from 0 to 10. Item scores were summed to yield a CDS total score ranging from 0 to 290, with scores above 70 indicating probable clinical DD.^46^ The CDS shows high internal consistency and good reliability^46^, and its factor structure has been previously analyzed in psychiatric samples.^47,48^

#### Anxiety and depression

State anxiety and depressive symptoms over the past seven days were assessed using the Hospital Anxiety and Depression Scale (HADS)^49^. It comprises 14 items rated on a 4-point scale, that form separate *anxiety* (HADS-A) and *depression* (HADS-D) factors. Scores ≥ 11 on either factor indicate probable disorder. Trait anxiety was assessed using the State-Trait Anxiety Inventory (STAI), Trait Form (Y-2)^50^, a 20-item questionnaire rated on a 4-point scale.

#### Impairment and symptom severity

Dizziness-related impairment was measured using the Dizziness Handicap Inventory (DHI)^51^, which contains 25 items rated on a 4-point scale. PPPD symptom severity was assessed using the French version of the Niigata PPPD questionnaire (NPQ)^11,52^, which contains 12 items rated on a 6-point scale. Total scores range from 0 to 72; scores ≥ 27 suggest a PPPD diagnosis.^52^

### Comparison of DD symptom severity with other clinical conditions

To contextualize the severity of DD symptoms observed in PPPD and OD, we systematically extracted mean CDS total scores, standard deviations, and proportion of individuals above the clinical cutoff of 70 from published studies that administered the 29-item CDS across psychiatric, functional and sensorimotor conditions (**Supplementary Table 1**).

### Statistical analyses

Analyses were carried out in JASP (version 0.19.3) and R (version 4.4.2).^53^ All tests were two-tailed with *P <* 0.05 denoting statistical significance unless specified otherwise. Sociodemographic and clinical variables were compared between groups using *χ*² tests, Mann−Whitney tests, and Kruskal−Wallis tests. Groups differences in DD, anxiety, depression, dizziness-related impairment, and PPPD symptom severity were assessed using Kruskal−Wallis tests with Bonferroni-corrected Dunn post-hoc tests.

To account for potential population-specific differences compared to prior CDS factor analyses^47,48^, we conducted an exploratory factor analysis (EFA) on all 29 items using the *psych* package with oblique promax rotation, given the expected intercorrelations between DD components.^47,48^ Only items with factor loadings >0.4 were retained. This analysis supported a four-factor solution (**Supplementary Tables 8-10**). A confirmatory factor analysis (CFA) was then performed using the *lavaan* package with the MLR estimator to evaluate configural, metric, and scalar invariance across groups. One factor was removed due to insufficient invariance. Scalar invariance was then achieved by releasing three intercept constraints (**Supplementary Table 11**). Internal consistency for retained factors was assessed using Cronbach’s α.

We tested the ability of DD factors, NPQ and DHI to discriminate PPPD from other OD, using logistic regressions and receiver operating characteristic (ROC) curve analyses. Area under the curve (AUC) values with 95% confidence intervals (CI) were estimated using stratified bootstrap resampling (2,000 replicates). AUC differences were compared with bootstrap, using the empirical distribution of AUC differences to derive 95% CIs and *p*-values. For patient stratification analyses, CDS, DHI, and NPQ scores were categorized into quantiles (quartiles and extreme deciles). Within each quantile and extreme decile, proportions of PPPD diagnosis were reported with 95% CI estimated using Wilson’s method. Logistic regression models using quantile categories (lowest = reference) and tests for linear trend were used to examine dose-response associations between questionnaire scores and subgroup membership.

Structural equation modelling (SEM; *lavaan* package) was employed to test whether anxiety and depression mediated the association between PPPD diagnosis (PPPD vs. OD) and DD symptoms. The model included CDS factors (measured by their respective items) identified by the EFA and CFA as latent endogenous variables, with depression, state anxiety, and trait anxiety as mediators. All CDS dimensions were assumed to be intercorrelated, as were depression and both state and trait anxiety. PPPD diagnosis served as exogenous variable (**Supplementary Fig. 1**). Age, sex, and migraine were included as covariates, given their established association with DD, anxiety, and depression^54–58^. SEM analyses used the Full Information Maximum Likelihood method to handle missing data, with 5,000 bootstrap repetitions for robust estimation.

### Data availability

The data that support the findings of this study are available from the corresponding author, upon reasonable request.

## Results

### Sample characteristics

The three groups were comparable in age (**Fig. 1b**), education, employment status, and smoking habits, but differed significantly in marital status and alcohol consumption. Migraine was more frequent in both patient groups than in HC (**Table 1**). In the PPPD group, 84% had a past or current otoneurological disorder and/or abnormal vestibular test results, while 33% were taking antidepressants at the time of inclusion. The distribution of diagnoses is shown in **Supplementary Table 2**.

PPPD patients had similar disease duration and migraine prevalence but showed less frequent peripheral vestibular and hearing impairment on available otoneurological testing compared with OD patients (**Supplementary Table 3**). Clinically, PPPD patients reported greater dizziness-related handicap (DHI: Median Difference (*MD*) = 16, *P <* 0.01) and higher symptoms severity (NPQ: *MD* = 8, *P <* 0.05) than OD patients (**Table 1**).

### Self-reported DD symptoms

CDS total scores varied significantly across groups (*H*(2) = 78.00, *P <* 0.001, *η*² = 0.17; **Fig. 2a**), with PPPD patients reporting substantially higher DD symptoms than OD patients (*MD* = 10, *P <* 0.01) and HC (*MD* = 23, *P <* 0.001). OD patients also scored higher than HC (*MD* = 13, *P <* 0.001). Similar patterns were observed for CDS frequency (*H*(2) = 71.51, *P <* 0.001, *η*² = 0.15) and duration (*H*(2) = 78.17, *P <* 0.001, *η*² = 0.17) (**Supplementary Tables 4−5**).

**Figure 2.**
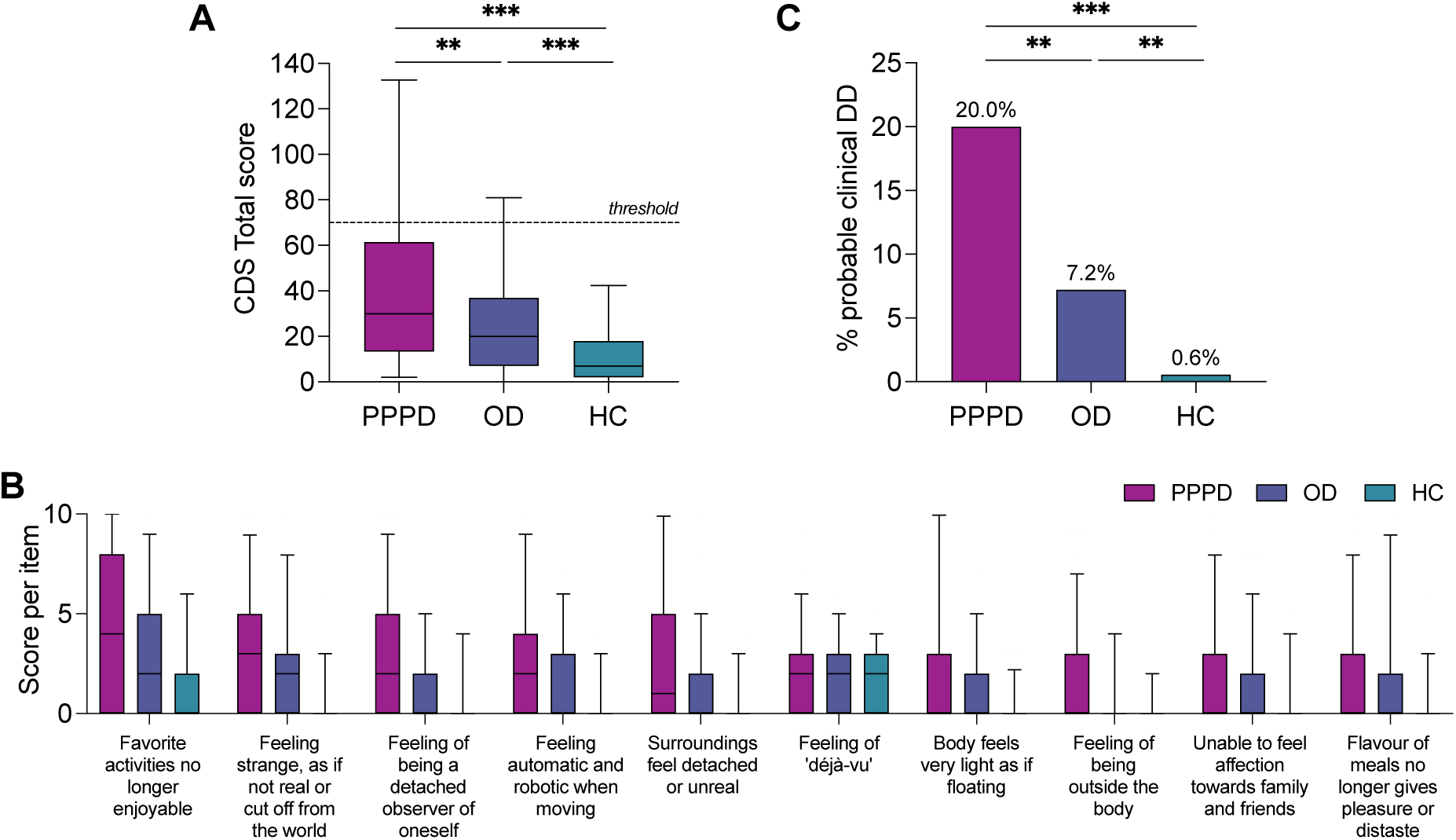
Depersonalization-derealization symptoms in persistent postural-perceptual dizziness. **(A)** Marked elevation of Cambridge Depersonalization Scale (CDS) total scores in persistent postural-perceptual dizziness (PPPD) compared with other otoneurological disorders (OD) and healthy controls (HC), shown as box-and-whisker plots (the solid line inside the box is the median, boxes cover the interquartile range, whiskers represent the 5^th^ to 95^th^ percentiles). Horizontal dashed line = clinical depersonalization-derealization threshold, i.e., CDS total score ≥ 70). Significant group differences: Bonferroni-corrected Dunn post-hoc tests (* corrected *P* < 0.05; ** corrected *P* < 0.01: *** corrected *P* < 0.001). **(B)** Symptom profile of the 10 highest-scoring CDS items in PPPD, illustrating the predominance of altered bodily- and self-related experiences. Scores are shown as box-and-whisker plots (same convention as in panel A). For full transcript of questionnaire items, see Supplementary Table 5. **(C)** Proportion of participants scoring above the clinical threshold, showing the higher occurrence of probable clinical depersonalization-derealization (DD) in PPPD patients. Bonferroni-corrected pairwise comparisons of proportions (* *P* < 0.05; ** *P* < 0.01; *** *P* < 0.001).

In PPPD patients, the most intense and/or frequently reported DD symptoms included depression (Q5), proper depersonalization (Q1), loss of first-person perspective (Q6), loss of agency (Q24), proper derealization (Q13), déjà-vécu (Q17), bodily lightness (Q8), out-of-body experience (Q23), affective detachment (Q16), and loss of taste-related pleasure (Q7) (**Fig. 2b**). Item-level analysis showed that PPPD patients scored significantly higher than OD patients on twelve items tapping into proper depersonalization and derealization, loss of body ownership, depression, first-person perspective alteration, bodily lightness, detachment from thoughts and voices, body image distortions, out-of-body experience and agency loss, and scored higher than HC on 18 items, while OD patients scored higher than HC on twelve items (*P*-adjusted < 0.05; **Supplementary Table 6−7**; **Supplementary Fig. 2**).

### Prevalence of probable DD disorder

Based on the CDS clinical cut-off of 70 (Ref^46^), the prevalence of probable clinical DD differed strongly between groups (*χ*²(2) = 34.77, *P <* 0.001), with rates of 20.0% in PPPD patients, 7.2% in OD patients, and 0.6% in HC (**Fig. 2c**). Logistic regression revealed that PPPD patients had a 3.2-fold higher odds of meeting the clinical DD threshold compared to OD patients (OR = 3.2, 95% CI [1.52, 6.78], *P <* 0.01) and a >40-fold higher odds compared to HC (OR = 43.50, 95% CI [5.74, 329.57], *P <* 0.001). OD patients also showed elevated risk compared to HC (OR = 13.55, 95% CI [1.75, 104.62], *P <* 0.05).

### Comparison with psychiatric, functional and sensorimotor disorders

PPPD patients exhibit DD symptom severity within the range reported for other forms of functional neurological disorders (e.g., psychogenic non-epileptic seizures), schizophrenia spectrum, and depressive or anxiety disorders. However, their symptom levels remained lower than those observed in trauma-related conditions (e.g., PTSD with childhood trauma), and well below those typically observed in primary dissociative disorders (e.g., DD disorder, dissociative identity disorder), which represent the extreme end of the CDS spectrum (**Fig. 3**). PPPD patients seem to experience more intense DD symptoms than those with sensorimotor conditions (e.g., other OD, spinal cord injury) or functional motor symptoms.

**Figure 3.**
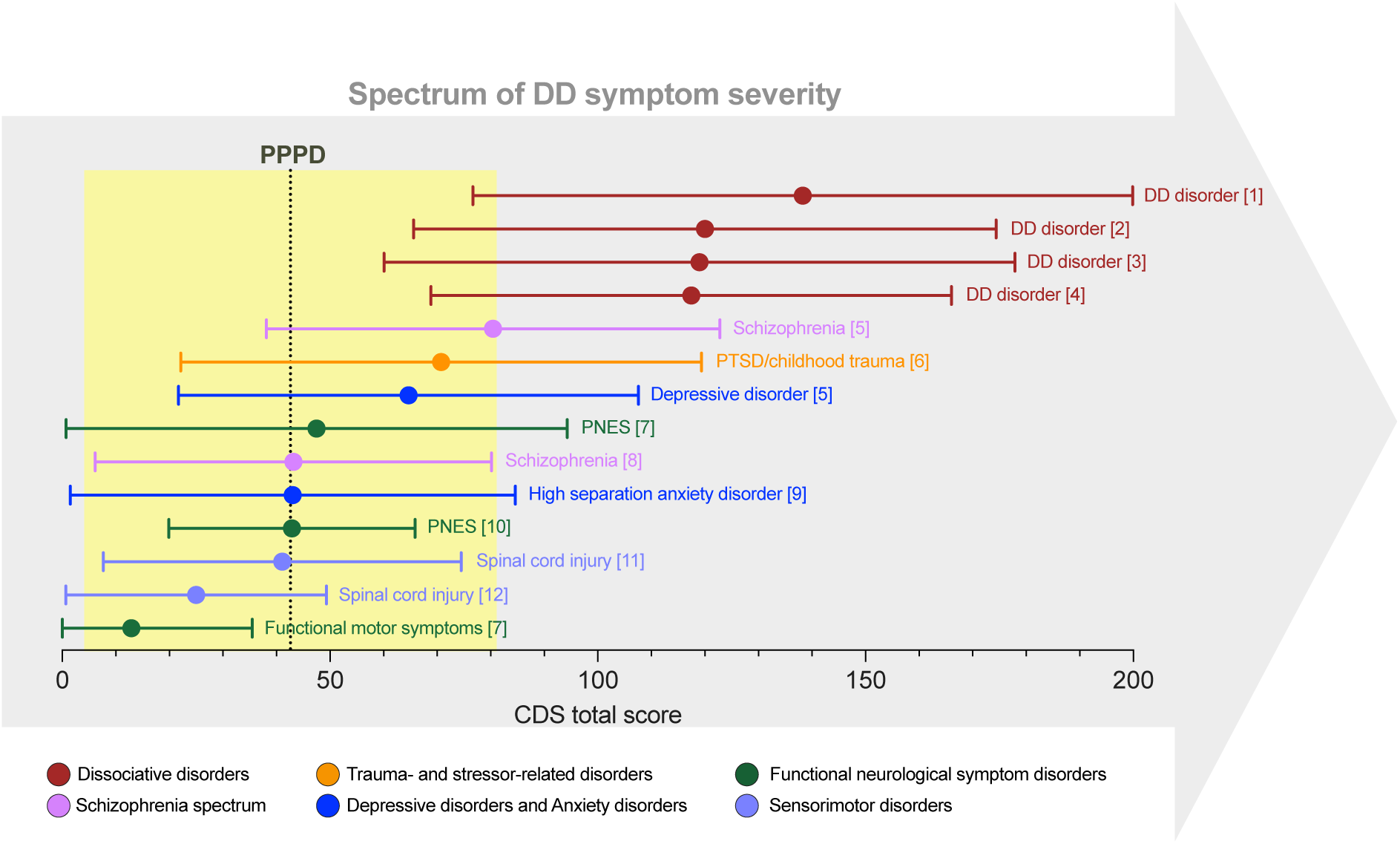
Spectrum of depersonalization-derealization symptoms severity in persistent postural-perceptual dizziness and various psychiatric and sensorimotor conditions. Mean Cambridge Depersonalization Scale (CDS) total scores (colored dots) and standard deviations (horizontal error bars) reported in published studies of various psychiatric and sensorimotor disorders (see Supplementary Table 1 for the list of studies [1−12]). Disorders are ordered by decreasing severity. Colors correspond to diagnostic groupings based on DSM-5-TR^85^ categories. The dotted vertical line and yellow shaded area represent the distribution (mean ± standard deviation) of CDS total scores observed in the present PPPD cohort. DD = depersonalization-derealization; PNES = psychogenic non-epileptic seizures; PTSD = post-traumatic stress disorder.

### Factor structure and group differences on the CDS

EFA and CFA revealed a three-factor structure for the CDS, including *Bodily Self Disturbances* (8 items)*, Cognitive and Affective Detachment* (7 items), and *Numbing* (3 items) (**Fig. 3a**), all factors showing good internal consistency (Cronbach’s *α* = 0.89, 0.80, and 0.76, respectively).

While significant group effects were observed for all CDS factors (all *P <* 0.05), only *Bodily Self Disturbances* clearly differentiated PPPD from OD patients (**Fig. 3b**; **Supplementary Tables 4-5**). This factor captured core bodily self alterations (body ownership, embodied self-location, agency, first-person perspective) and showed the most robust group difference (*H*(2) = 90.84, *P <* 0.001, *η*² = 0.20). PPPD patients scored higher than both OD patients (*MD* = 1.00, *P <* 0.001) and HC (*MD* = 1.75, *P <* 0.001), while OD patients also scored higher than HC (*MD* = 0.75, *P <* 0.001). For *Cognitive and Affective Detachment*, only a smaller group effect emerged (*H*(2) = 6.45, *P <* 0.05, *η*² = 0.00), driven by a minimal difference between PPPD patients and HC (*MD* = 0.07, *P <* 0.05). OD patients did not differ from either group (*P* ≥ 0.230). Finally, *Numbing* showed a moderate group effect (*H*(2) = 33.09, *P <* 0.001, *η*² = 0.07). Both PPPD (*MD* = 0.00, *P <* 0.001) and OD patients (*MD* = 0.00, *P <* 0.001) scored higher than HC, but did not differ from one another (*MD* = 0.00, *P =* 0.535).

**Figure 4.**
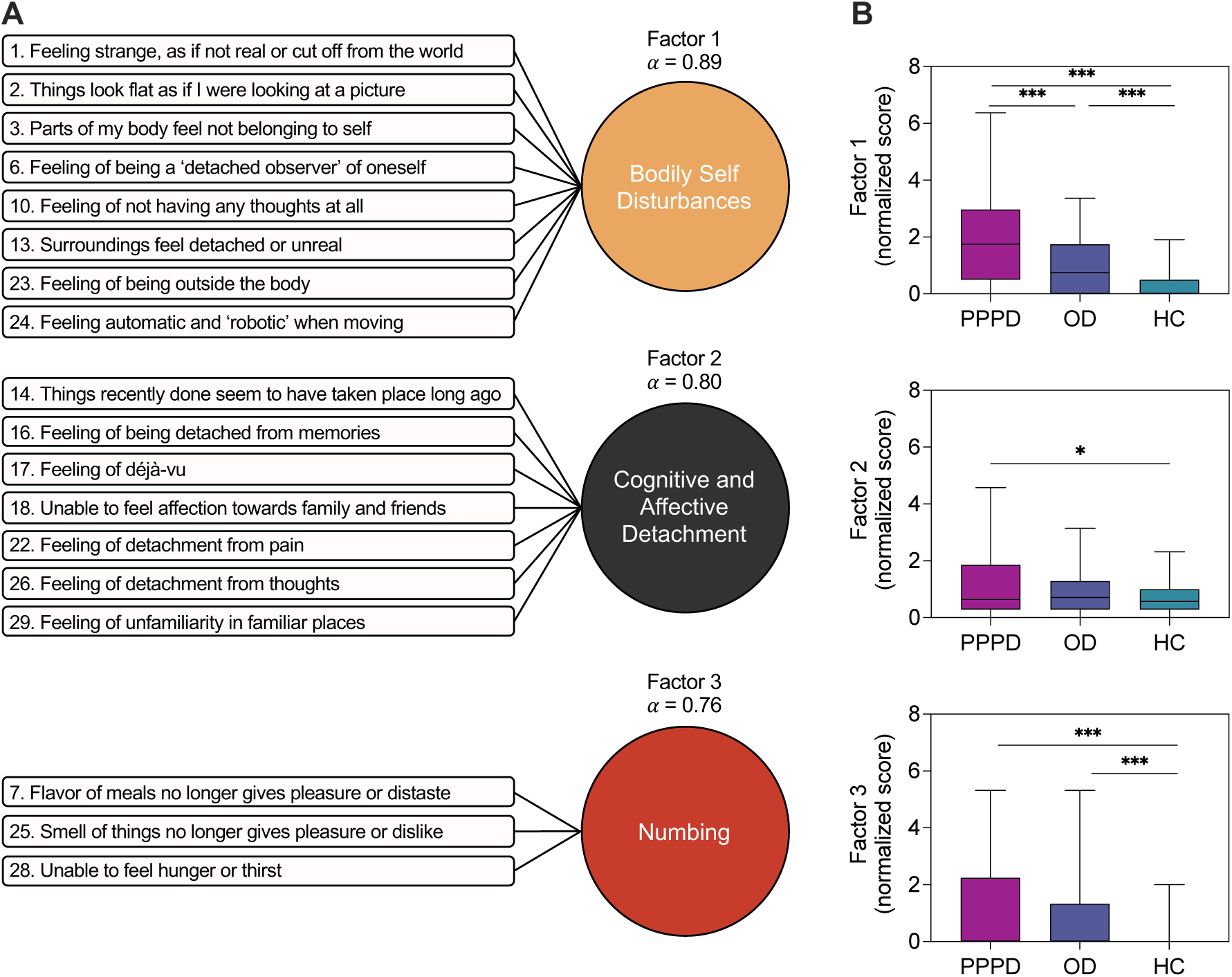
Factor structure of the Cambridge Depersonalization Scale. **(A)** Three-factor solution of the Cambridge Depersonalization Scale (CDS) showing item loadings on each factor and internal consistency (Cronbach’s *α*). **(B)** Group differences in normalized factor scores, shown as box-and-whisker plots (the solid line inside the box is the median, boxes cover the interquartile range, whiskers represent the 5^th^ to 95^th^ percentiles). Scores were normalized by dividing each factor score by its number of items, enabling comparison across factors. Significant group differences: Bonferroni-corrected Dunn post-hoc tests (* corrected *P* < 0.05; ** corrected *P* < 0.01: *** corrected *P* < 0.001).

### Predictive value of DD symptoms

Of all CDS factors, only *Bodily Self Disturbances* significantly predicted PPPD diagnosis (OR = 1.404, 95% CI [1.206–1.657], *P <* 0.001), with moderate sensitivity (0.49) and good specificity (0.77) (**Fig. 5a**). ROC analyses showed that *Bodily Self Disturbances* discriminated PPPD from other OD better than chance (AUC = 0.659, 95% CI [0.588–0.726], *P <* 0.001), whereas the other factors did not (*Cognitive and Affective Detachment*: AUC = 0.536, *P =* 0.148; *Numbing*: AUC = 0.543, *P =* 0.082). *Bodily Self Disturbances* performed as good as established clinical measures such as DHI (AUC = 0.674, *P <* 0.001; AUC difference = −0.016, *P =* 0.650) and NPQ (AUC = 0.608, *P <* 0.001; AUC difference = 0.049, *P =* 0.177) (**Fig. 5b; Supplementary Tables 12−13**).

**Figure 5.**
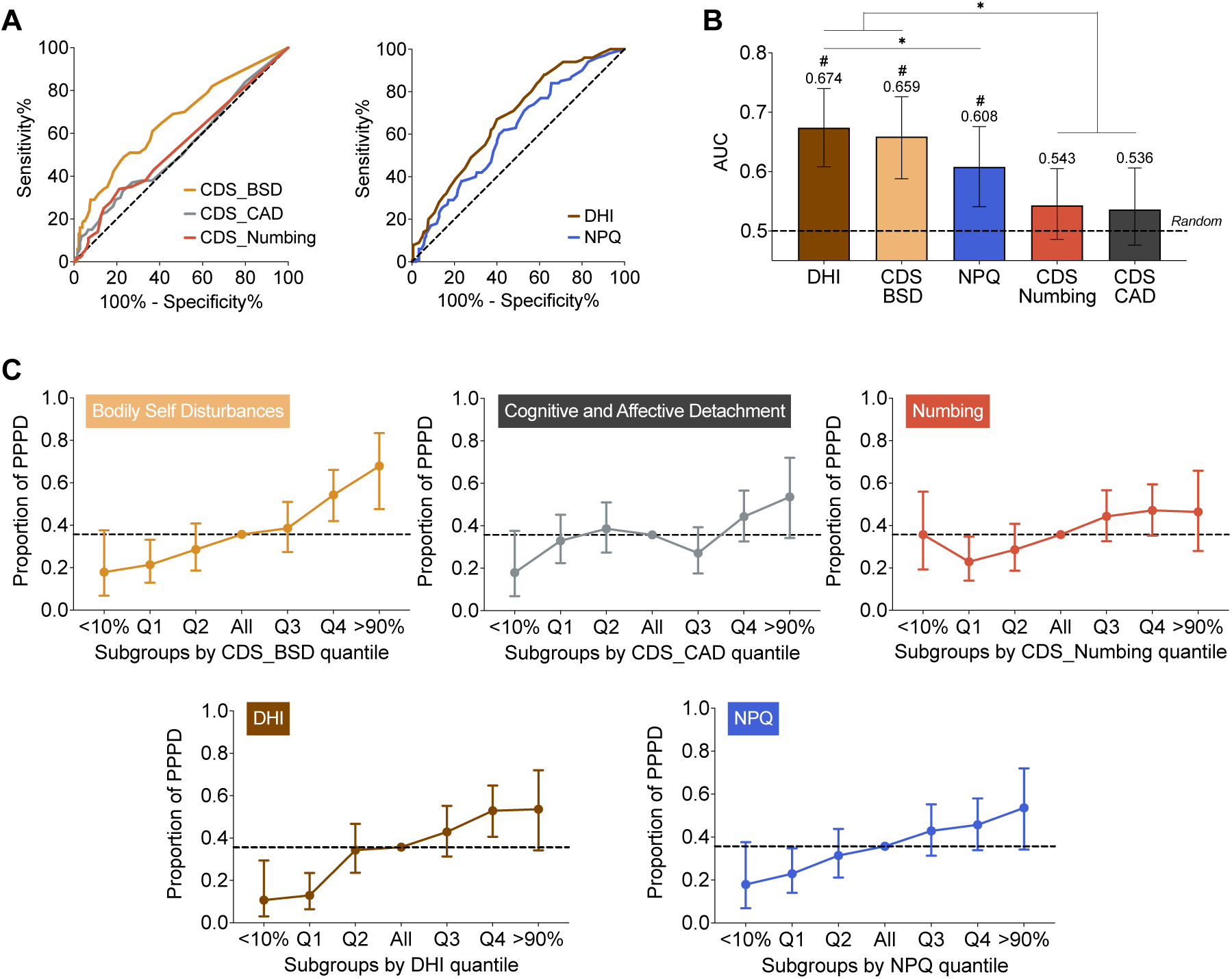
Discriminative performance of Cambridge Depersonalization Scale factors and symptom questionnaires for persistent postural-perceptual dizziness. **(A)** Receiver operating characteristic (ROC) curves for the three Cambridge Depersonalization Scale (CDS) factors (BSD = *Bodily Self Disturbances*; CAD = *Cognitive and Affective Detachment*; *Numbing*), Dizziness Handicap Inventory (DHI), and Niigata PPPD Questionnaire (NPQ). **(B)** Area under the ROC curve (AUC) for each measure, showing the relative discriminative ability of the questionnaires (dashed line = chance level, i.e., AUC = 0.5). # indicates significant above chance level discrimination (*P* < 0.05). * indicates significant differences between AUC (bootstrap comparison of ROC curves; *P* < 0.05). **(C)** Proportion of PPPD diagnoses across strata (extreme deciles and quartiles) for each questionnaire (error bars: 95% CI; dashed line: overall proportion of PPPD diagnosis).

Quantile analysis further confirmed the discriminative ability of *Bodily Self Disturbances* (**Fig. 5c; Supplementary Tables 14−15**). The likelihood of PPPD diagnosis increased progressively with higher *Bodily Self Disturbances* scores: participants in the third quartile were more than twice as likely to have PPPD as those in the first quartile (OR = 2.30, 95% CI [1.10–4.95], *P <* 0.05), and those in the fourth quartile were over four times as likely (OR = 4.35, 95% CI [2.11–9.34], *P <* 0.001). The effect was particularly pronounced at the extremes: participants in the top 10 % of *Bodily Self Disturbances* scores had nearly tenfold higher odds of PPPD compared with the lowest decile (OR = 9.70, 95% CI [2.96–37.16], *P <* 0.001).

Logistic regression per quartiles confirmed a significant trend for *Bodily Self Disturbances* (OR per quartile = 1.638, 95% CI [1.30–2.08], *P <* 0.001). In contrast, *Cognitive and Affective Detachment* showed no significant increase across quartiles (OR per quartile = 1.11, 95% CI [0.89–1.38], *P =* 0.373), and *Numbing* showed only a modest association (OR per quartile = 1.49, 95% CI [1.19–1.88], *P <* 0.01). Odds of PPPD also increased with higher DHI scores (OR per quartile = 1.813, 95% CI [1.43–2.32], *P <* 0.001) and NPQ scores (OR per quartile = 1.428, 95% CI [1.14–1.80], *P <* 0.01).

### Self-reported anxiety and depression

Robust group effects were found for trait anxiety (*H*(2) = 50.41, *P <* 0.001, *η*² = 0.11), state anxiety (*H*(2) = 36.63, *P <* 0.001, *η*² = 0.08), and depression (*H*(2) = 68.24, *P <* 0.001, *η*² = 0.15). Both patient groups scored higher than HC on trait anxiety (PPPD *vs* HC: *MD* = 12, *P <* 0.001; OD *vs* HC: *MD* = 7, *P <* 0.001), state anxiety (PPPD *vs* HC: *MD* = 4, *P <* 0.001; OD *vs* HC: *MD* = 2, *P <* 0.001), and depression (PPPD *vs* HC: *MD* = 4, *P <* 0.001; OD *vs* HC: *MD* = 2, *P <* 0.001) (**Fig. 6a**). PPPD patients reported higher trait anxiety (MD = 5, *P <* 0.05) and depression (*MD* = 2, *P <* 0.001), than OD patients, whereas state anxiety did not differ significantly (*P =* 0.07).

**Figure 6.**
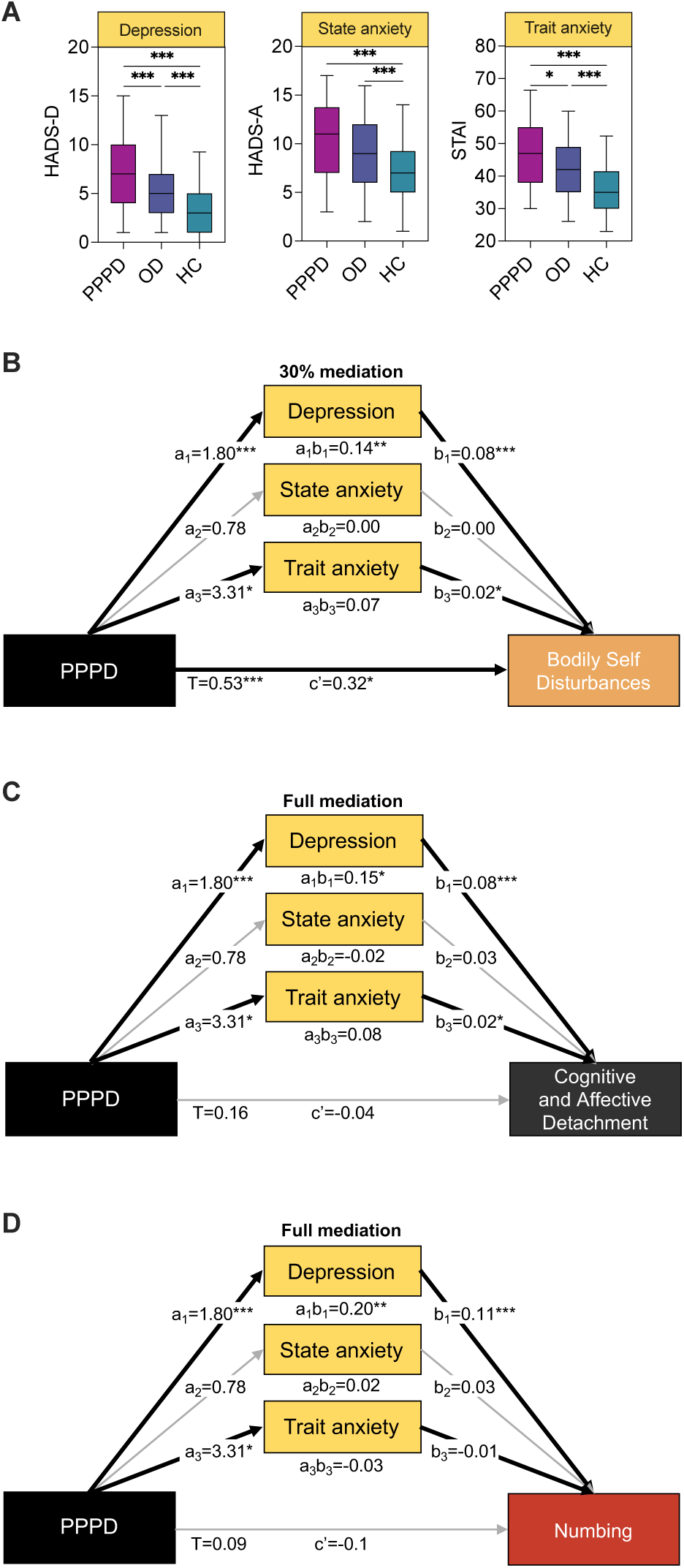
Mediation of the effect of persistent postural-perceptual dizziness on depersonalization-derealization component by depression and anxiety. **(A)** Box-and-whisker plots showing depressive symptoms (HADS-D), state anxiety (HADS-A) and trait anxiety (STAI) in patients with persistent postural-perceptual dizziness (PPPD), other otoneurological disorders (OD), and healthy controls (HC). The solid line inside the box is the median, boxes cover the interquartile range, whiskers represent the 5^th^ to 95^th^ percentiles. Significant group differences: Bonferroni-corrected Dunn post-hoc tests (* corrected *P* < 0.05; ** corrected *P* < 0.01: *** corrected *P* < 0.001). **(B–D)** Structural equation models testing whether depression, state anxiety and trait anxiety mediate the association between PPPD and each CDS factor: (B) *Bodily Self Disturbances*, (C) *Cognitive and Affective Detachment*, and (D) *Numbing*. Standardized beta values for latent variables are reported. Direct effects are denoted c’; indirect effects reflect the product of paths for depression (a_1_b_1_ = a_1✕_b_1_), state anxiety (a_2_b_2_ = a_2✕_b_2_), and trait anxiety (a_3_b_3_ = a_3✕_b_3_). Total effects (T) are the sum of direct and indirect effects (T = c’ + a_1_b_1_ + a_2_b_2_ + a_3_b_3_). Percent mediation refers to the proportion of the total effect explained by significant indirect pathways and is calculated as a_x_b_x_/T. Inconsistent mediations are considered as full mediation, with no computation of mediation proportion. Covariates (age, sex, migraine), covariances between depression and anxiety, and covariances between CDS subscales are included in the models but omitted from the graphs for clarity. Significant paths: * *P* < 0.05; ** *P* < 0.01: *** *P* < 0.001.

### Mediation of DD by anxiety and depression

The structural equation model showed good fit to the data (**Supplementary Table 16**).

For *Bodily Self Disturbances*, PPPD diagnosis (compared with OD) exhibited a large total effect (*β(total)* = 0.53, 95% CI [0.26, 0.79], *P <* 0.001). Importantly, this relationship persisted after controlling for depression, anxiety, age, sex, and migraine history, with a significant direct effect of PPPD on *Bodily Self Disturbances* (*β(direct)* = 0.32, 95% CI [0.06, 0.58], *P <* 0.01). Depression partially mediated this relationship (*β(indirect)* = 0.14, 95% CI [0.04, 0.25], *P <* 0.01), explaining 27.4% of the total effect. By contrast, neither state (*P =* 0.959) nor trait anxiety (*P =* 0.099) mediated the relationship. Age showed a small negative effect (*β* = –0.01, 95% CI [–0.02, –0.00], *P <* 0.01), whereas migraine and sex had no significant effect (**Fig. 6b**; **Supplementary Tables 17−18**).

For *Cognitive and Affective Detachment*, PPPD had no direct (*P =* 0.733) or total (*P =* 0.296) effect. The association with PPPD was fully mediated by depression (*β(indirect)* = 0.15, 95% CI [0.04, 0.27], *P <* 0.01). State (*P =* 0.319) and trait anxiety (*P =* 0.073) were non-significant mediators; covariates had no significant effect (**Fig. 6c**; **Supplementary Tables 17−18**).

Similarly, for *Numbing*, PPPD showed no direct (*P =* 0.464) or total (*P =* 0.094) effects. The effect of PPPD was entirely mediated by depression (*β(indirect)* = 0.20, 95% CI [0.07, 0.34], *P <* 0.001), with no mediation through state (*P =* 0.353) or trait anxiety (*P =* 0.359) and no significant contribution of covariates (**Fig. 6d**; **Supplementary Tables 17−18**).

## Discussion

This study provides a novel, comprehensive examination of DD symptoms in PPPD, exploring their relationship with anxiety and depression in a large cohort of 455 participants. Key findings reveal that PPPD patients exhibited significantly higher DD scores than both HC and patients with other OD. The prevalence of probable clinical DD was markedly elevated in PPPD (20%), compared to 7.2% in OD and 0.6% in HC. While DD severity in PPPD was below levels observed in dissociative disorders, it was comparable to that of functional neurological disorders. Factor analysis identified three distinct DD dimensions: *Bodily Self Disturbances*, *Cognitive and Affective Detachment*, and *Numbing*. Only *Bodily Self Disturbances* significantly differentiated PPPD from other OD, demonstrating robust classification performance (AUC = 0.659) comparable to established clinical tools (DHI, NPQ). *Bodily Self Disturbances* was the only DD factor directly associated with PPPD, independent of psychological factors, age, sex, and migraine. Importantly, depression partially mediated the relationship between PPPD and *Bodily Self Disturbances* and fully mediated the relationship for *Cognitive and Affective Detachment* and *Numbing*, while anxiety had no mediating effect. These findings not only refine the phenotyping of PPPD but also highlight bodily self disturbances and depression as potential targets for diagnostic and therapeutic innovation.

### Symptoms of DD are more intense in PPPD than in OD

This study provides the first large-scale characterization of DD symptoms in PPPD, demonstrating that DD symptoms are significantly more intense and frequent in PPPD compared to both individuals with other OD and HC. While DD symptoms have been described in OD, where they are more pronounced than in the general population^35–38^, their systematic evaluation in PPPD has been limited. Previous work included a qualitative study reporting DD experiences in a small PPPD sample^44^, and a quantitative study using the Depersonalization and Derealization Inventory (DDI)^59^, which found higher DD scores than in healthy individuals. Using the CDS, a validated and reliable instrument assessing a large spectrum of DD symptoms^46–48^, our findings extend these observations. We show that DD symptoms in PPPD not only exceed those observed in healthy individuals but are also more severe than in other forms of OD.

In our cohort, 84% of PPPD patients presented with secondary PPPD, characterized by a history of vestibular impairment. Comorbid vestibular abnormalities are common in PPPD, especially in tertiary vertigo centres.^3,6,8^ Yet, we found that DD symptoms were more pronounced in PPPD patients despite milder vestibular and hearing impairment compared to OD patients. This observation aligns with clinical evidence indicating that the degree of vestibular deficit is a weak predictor of PPPD development^7^. Thus, DD symptoms in PPPD likely reflect disorder-specific mechanisms rather than mere vestibular dysfunction.

The association of migraine with both PPPD^1,60^ and DD^56^ could theoretically contribute to the observed relationship, but PPPD patients exhibited higher DD scores despite comparable migraine prevalence relative to other OD. Furthermore, migraine had no significant effect on DD factors in the structural equation models, suggesting that its role in DD symptoms is limited. This reinforces the hypothesis that DD symptoms in PPPD are primarily driven by mechanisms intrinsic to the disorder itself, rather than by comorbid conditions such as migraine or vestibular impairment.

### Prevalence and severity of clinical DD in PPPD

Twenty percent of PPPD patients met the CDS threshold for probable clinical DD, representing the first large-scale estimate of clinical DD prevalence in this population. This rate was threefold higher than in OD patients and more than thirtyfold higher than in HC. While transient DD experiences are common in the general population (9.7–74% of individuals)^32,54^, only 1–2% meet criteria for clinical DD.^31,54^ Thus, the prevalence observed in our PPPD cohort is markedly elevated relative to both the general population and other OD. This prevalence aligns with rates reported in patients with visual snow syndrome^61^, mood or anxiety disorders (∼3–20%)^31,62^, and schizophrenia (17%)^63^, as assessed using the same psychometric instrument (CDS)^46^. However, it remains well below the prevalence observed in primary dissociative disorders (79%).^48^ This suggests that DD symptoms in PPPD, while clinically significant, do not reach the severity seen in dissociative pathology.

In terms of severity (CDS total score, **Fig. 3**), DD symptoms in PPPD did not reach the levels typically observed in primary dissociative disorders but fell within the range reported for functional neurological disorders (e.g., psychogenic non-epileptic seizures) and some psychiatric conditions (e.g., schizophrenia)^34^. These findings support the conceptualization of PPPD within the spectrum of functional neurological disorders^6,19^. PPPD shares key clinical and pathophysiological features with functional neurological disorders, including mechanisms explaining symptom triggering and chronicity. Both disorder types have been interpreted using the predictive processing framework^19,64^ (see below). In addition, functional neurological disorders are intrinsically linked to dissociative symptoms and bodily self disturbances^19,23,65,66^, sometimes associated with reduced interoceptive acuity^67^. Notably, functional neurological disorders involve a loss of agency over the body^19^, a feature that aligns with disruptions in agency for movements and thoughts observed in our PPPD cohort.

The comparison of DD prevalence and severity across clinical conditions indicates that DD symptoms in PPPD are not marginal. Instead, they reach levels comparable to those observed in functional and psychiatric disorders. Importantly, the elevation of DD symptoms in PPPD cannot be attributed to vestibular impairment alone, as comparable DD levels are not observed in OD patients with more severe vestibular deficits. Together, these findings highlight the clinical distinctiveness of PPPD and indicate that DD symptoms represent a significant dimension of its phenomenology.

### Bodily self disturbances: a specific marker of PPPD

Factor analysis on DD symptoms resulted in three factors: *Bodily Self Disturbances*, *Cognitive and Affective Detachment*, and *Numbing*. Among them, only *Bodily Self Disturbances* differed significantly between PPPD and OD, which aligns with the most frequently and severely reported items in PPPD patients. This factor encompasses alterations in core phenomenal dimensions of the bodily self, including body ownership, first-person perspective, self-location, and agency, concepts well-established in recent neuroscientific literature^25^.

Bodily self disturbances have been described in patients with other OD, including Alice in Wonderland syndrome and out-of-body experiences in vestibular migraine and peripheral vestibular disorders^39–41^, and some of them can even be experimentally induced in healthy participants through caloric vestibular stimulation.^35,40^ However, these disturbances appear substantially more pronounced in PPPD, where they had not been systematically investigated prior to this study.

Interestingly, the two other DD factors, *Cognitive and Affective Detachment* (detachment from thoughts, emotions, and memories) and *Numbing* (loss of emotional coloring for taste and smell) were similarly intense in PPPD and OD. These symptoms may stem from attenuated emotional and autonomic responses to vertigo and vestibular dysfunction^68,69^. In addition, they may arise from cognitive resource reallocation towards balance and posture monitoring^36^, which could limit resource availability for processing memory, thoughts and emotions. However, our results suggest that *Cognitive and Affective Detachment* and *Numbing* are fully mediated by depression, which may explain the lack of differences between PPPD and OD on these factors (see below).

Importantly, *Bodily Self Disturbances* was also the only DD factor that reliably discriminated PPPD from other OD. Its classification accuracy was comparable to that of established tools such as the DHI, commonly used to quantify dizziness-related impairment, and to the NPQ, which was developed based on PPPD diagnostic criteria. While the NPQ typically shows moderate discriminative performance (AUC: 0.661− 0.780)^11,52,70,71^, its accuracy was lower in our sample (AUC: 0.608). In contrast, the *Bodily Self Disturbances* factor achieved an AUC of 0.659, which was numerically higher than that of the NPQ, demonstrating its potential as a clinical marker of PPPD.

In addition, we observed a strong dose-response relationship between *Bodily Self Disturbances* and PPPD diagnosis. Specifically, 67.9% of patients in the highest decile of *Bodily Self Disturbances* had a PPPD diagnosis, compared to 54.3% in the highest quartile. Patients in the top decile demonstrated nearly tenfold higher odds of PPPD relative to the lowest decile, while patients in the highest quartile had more than fourfold higher odds. This dose-response relationship was comparable to that observed for the NPQ and DHI, but significantly stronger than for *Cognitive and Affective Detachment* and *Numbing*.

In conclusion, the specificity of PPPD does not lie in general DD symptoms, but rather in *Bodily Self Disturbances*. This factor not only distinctly characterizes PPPD but also differentiates it from other otoneurological disorders. Unlike the other DD factors, *Bodily Self Disturbances* demonstrates robust classification abilities. Therefore, we propose that it constitutes a reliable clinical marker of PPPD, with significant potential for implementation in clinical practice.

### Only bodily self disturbances relate to PPPD beyond anxiety and depression

Structural equation modelling revealed that *Bodily Self Disturbances* was specifically associated with PPPD, even after accounting for the mediating effects of anxiety and depression. In contrast, other DD dimensions were fully mediated by depression. This indicates that, beyond psychological factors, age, sex, and migraine, PPPD shows a unique and direct relationship with *Bodily Self Disturbances*.

In OD, bodily self disturbances have been interpreted as a consequence of multisensory mismatch caused by vestibular impairment, leading to distortions in body and environment representations.^36,39,42^ However, in PPPD, the intensity and specificity of *Bodily Self Disturbances* suggest mechanisms that extend beyond a mere sensory conflict.

Current pathophysiological models of PPPD emphasize altered postural control strategies, excessive visual dependence, heightened motion sensitivity, and increased conscious monitoring of posture^1^. These processes are associated with an overestimation of postural sway^72^ and altered spatial processing^73^, which have been related to alterations in multisensory integration^10,16,17^ and heightened sensitivity to multisensory conflict^18^. Neuroimaging studies support this view, revealing structural, functional and connectivity alterations in key multisensory regions, including the parieto-insular vestibular cortex, superior temporal gyrus, hippocampus, and supramarginal gyrus.^16,17^ As these regions are part of the brain network underpinning bodily self dimensions^25,74^, their disturbances could account not only for the core symptoms of PPPD but also for bodily self disturbances observed in this study.

PPPD and other functional neurological disorders have been conceptualized within the predictive processing framework, which posits that they result from erroneous and overweighted top-down priors^1,19,75^, shaped by previous expectations and emotional salience, associated with a reduced weighting of bodily signals.^19,23^ A recent formulation suggests that PPPD involves a failure in sensory attenuation.^64^ In this model, cognitive beliefs of imbalance reduce the weighting of self-motion priors, while prediction errors for vestibular afferences are overweighted. Thus, normally fluctuating vestibular input is not adequately attenuated during self-motion, causing a shift from a top-down ‘autopilot’ to a bottom-up ‘feedback’ mode. This shift results in subjective postural instability and may underlie the bodily self disturbances observed in PPPD. Within this framework, altered priors concerning the body and its movements, or disrupted sensory attenuation, combined with hypervigilance towards bodily sensations, could compromise the normally pre-reflexive and transparent nature of bodily self dimensions.^76^ This interpretation is consistent with theoretical accounts of DD symptoms as arising from a failure in sensory attenuation and the consequent loss of transparency of self-referential processes.^24^

Finally, it is conceivable that bodily self alterations may play a predisposing role in the development of PPPD. Both PPPD and DD symptoms have been proposed to lie on a spectrum across healthy individuals and clinical populations.^15,77^ Altered body representations may therefore represent a shared vulnerability factor for motion hypersensitivity and bodily self disturbances, thereby enhancing vulnerability to PPPD. Future longitudinal studies are required to determine whether such alterations precede PPPD onset or emerge as consequences of the disorder.

### Role of depression in mediating DD symptoms

In this study, trait anxiety was significantly higher in PPPD patients compared to those with other OD, whereas state anxiety did not differ between the two groups. This aligns with pathophysiological models of PPPD, which emphasize anxiety-related traits as predisposing factors for the disorder.^1,7^ In contrast, state anxiety appears to be more closely related to dizziness in general and does not consistently distinguish PPPD from OD.^11,12,73^ However, in our results, neither trait nor state anxiety mediated the relationship between PPPD and DD symptoms, in line with mixed findings in the literature regarding the association between anxiety and DD^78^. It is also possible that our measures of anxiety and depression overlap, with depression capturing the shared variance between these psychological constructs.

By contrast, depression was significantly higher in PPPD than in both OD and HC. Although often overlooked, depressive symptoms have been reported in PPPD patients^11,59,79^, associated with greater dizziness-related handicap and reduced quality of life.^79^ These symptoms may result from the negative impact of PPPD on employment, social relationships, leisure and daily activities^79^. Additionally, depressive symptoms may stem from reciprocal connections between the vestibular system and neural networks involved in emotion processing^80^. Furthermore, PPPD and depression share core mechanisms such as heightened body and symptom vigilance and negative interpretations of bodily sensations, which contribute to rumination and symptom perpetuation.^1,81^

Importantly, we show that depression emerged as a strong mediator of DD symptoms in PPPD. Depression fully mediated the relationship between PPPD and *Cognitive and Affective Detachment* as well as *Numbing*, while it partially mediated the relationship with *Bodily Self Disturbances*. This is consistent with evidence that DD disorders are highly comorbid with depressive disorders.^31,55,82^ Depressive processes, such as rumination, withdrawal, and thought suppression regarding the self have been related to DD experiences, particularly detachment and numbing.^83^ This may account for the complete mediation of these DD dimensions by depression. Regarding bodily self disturbances, depressive processes may exacerbate disruptions in bodily self transparency by enhancing attention to overweighted priors or sensory prediction errors.^24,64^

These findings highlight depression as a major perpetuating factor for both PPPD and DD symptoms, with important implications for clinical management. Systematic assessment and management of depressive symptoms may therefore be crucial for alleviating DD symptoms and improving overall management of PPPD. The prominent role of depression may also explain the clinical efficacy of selective serotonin reuptake inhibitors in PPPD, alongside their effects on serotonergic modulation within the vestibular-emotional neural networks.^84^

## Limitations

Several limitations should be acknowledged. First, vestibular function was not assessed in the HC group, as inclusion was based on the absence of any history of vertigo or dizziness. However, the presence of presbyvestibulopathy, characterized by reduced vestibular reflexes, is unlikely in our HC cohort, given that none reported vestibular symptoms, unsteadiness, gait disturbances, or falls. Second, the cross-sectional design limits the strength of causal inferences. While our statistical models are compatible with a directional relationship from PPPD to DD symptoms, reverse or bidirectional relationships cannot be ruled out. Longitudinal studies are needed to determine whether PPPD precedes DD symptoms, whether DD symptoms contribute to PPPD development, or if these phenomena interact dynamically over time. Third, DD symptoms were assessed using the CDS, which evaluates them over the past 6 months, whereas most patients in our sample had a longer disease duration. Yet, DD symptoms may fluctuate over time and according to disease state, and transient exacerbations of DD severity may occur during vertigo episodes. Finally, most patients in our sample presented with secondary PPPD and a history of vestibular dysfunction, reflecting recruitment from a tertiary vertigo center, where secondary PPPD is more prevalent than in general neurology clinics.^8^ This may limit the generalizability of our findings to patients with primary PPPD or to those without vestibular comorbidity.

## Conclusions

This study provides evidence that bodily self disturbances represent the most distinctive dimension of DD in PPPD, distinguishing it from other common OD. Our findings advance the clinical phenotyping of PPPD by identifying a potential marker of the disorder. We propose that the *Bodily Self Disturbances* factor of the CDS could serve as a useful clinical tool to detect these alterations in patients with PPPD and, potentially, identify individuals at risk of developing PPPD.

In addition, our results highlight the significant contribution of depressive symptoms to DD in PPPD, a factor that has received limited attention in the literature. These findings support a systematic assessment and management of depressive symptoms in PPPD, which may improve both symptom control and quality of life.

More broadly, this work refines current pathophysiological models of PPPD by identifying a core phenomenological feature, bodily self disturbance, and by elucidating its relationship with psychological comorbidities, particularly depression.

## Acknowledgements

We thank Dr Maeva Montero and the staff of the *Centre des Vertiges* at the European Hospital-Marseille, as well as the Clinical Research Department of the European Hospital-Marseille (the clinical study sponsor), for their invaluable assistance with patient inclusion and testing. We are grateful to Marco Bressan for his advice on structural equation modelling.

## Supplementary material

Supplementary material is available at *Brain* online.

## Supplementary Methods

**Supplementary Table 1.**
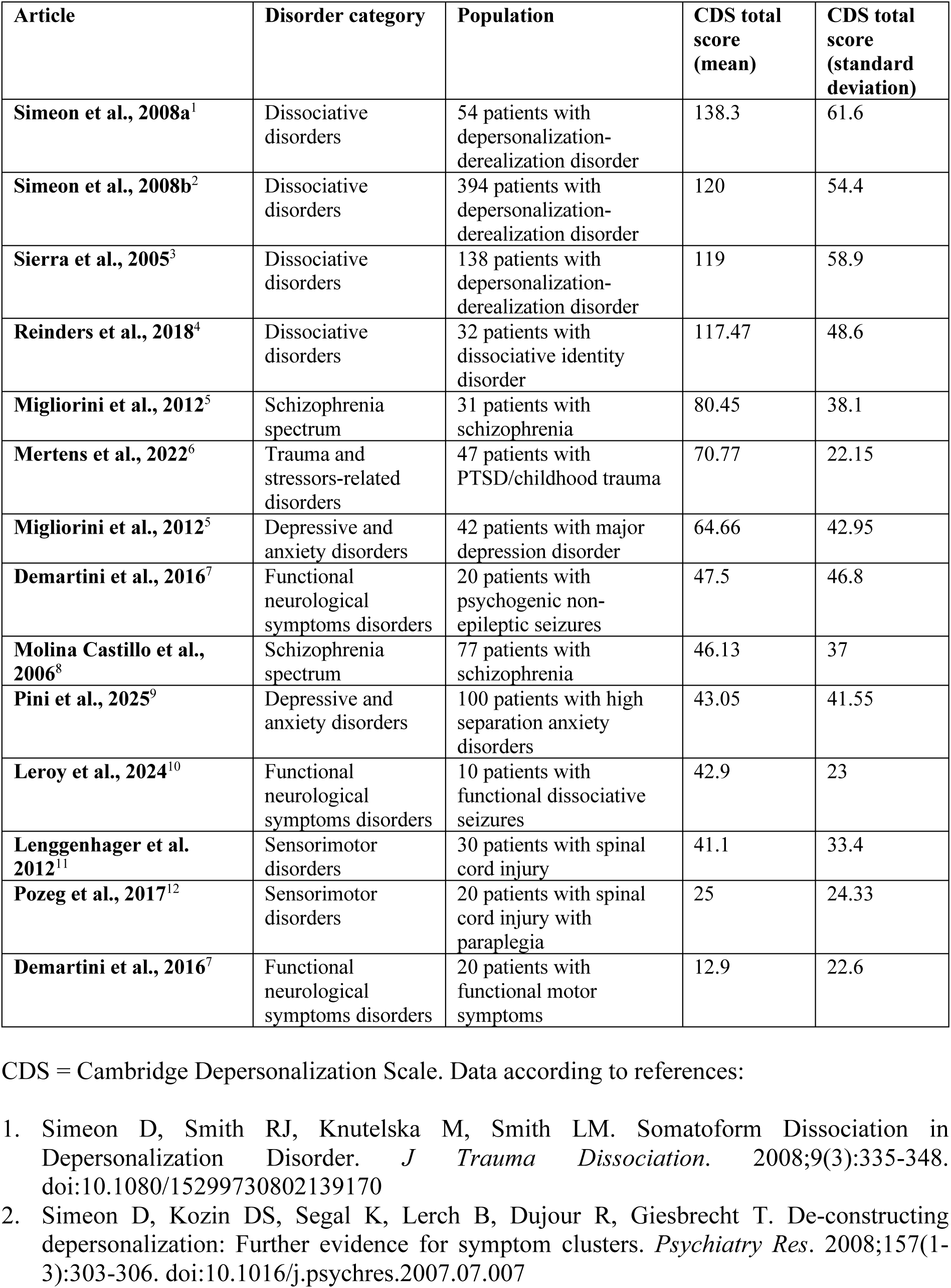
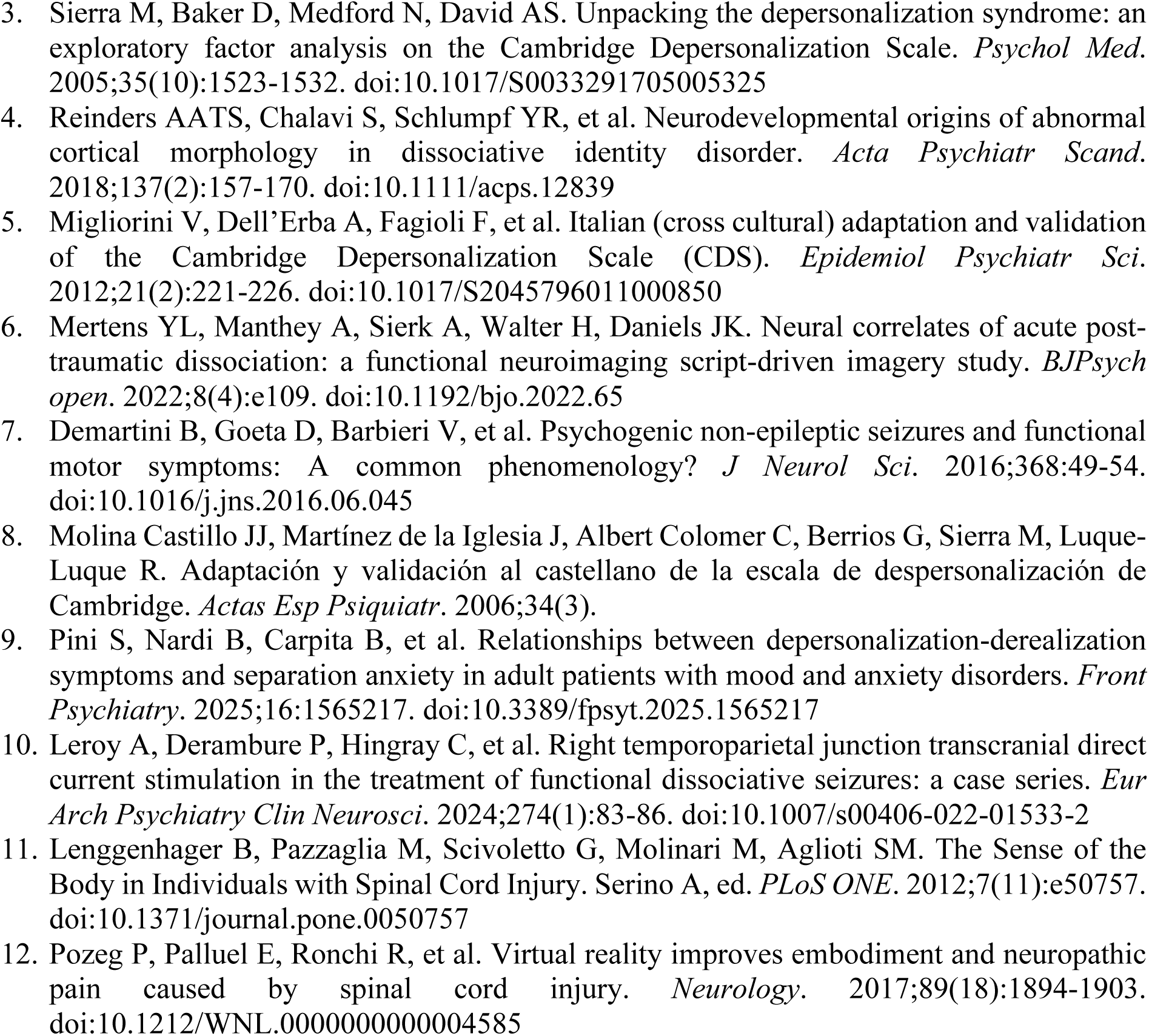
Summary of studies reporting depersonalization-derealization symptoms severity in various psychiatric and sensorimotor conditions.

## Supplementary Methods

### Structural equation modelling and mediation analysis

In the mediation models, indirect effects were calculated as the product of the coefficients forming each mediator’s pathway. The total effect corresponds to the sum of the direct and indirect effects. The proportion mediated was computed by dividing the indirect effect by the total effect. When mediation was inconsistent, i.e., when the direct effect was negative and the indirect effect exceeded the total effect, we considered the mediation to be full and did not compute a mediation proportion^13,14^.

**Supplementary Figure 1.**
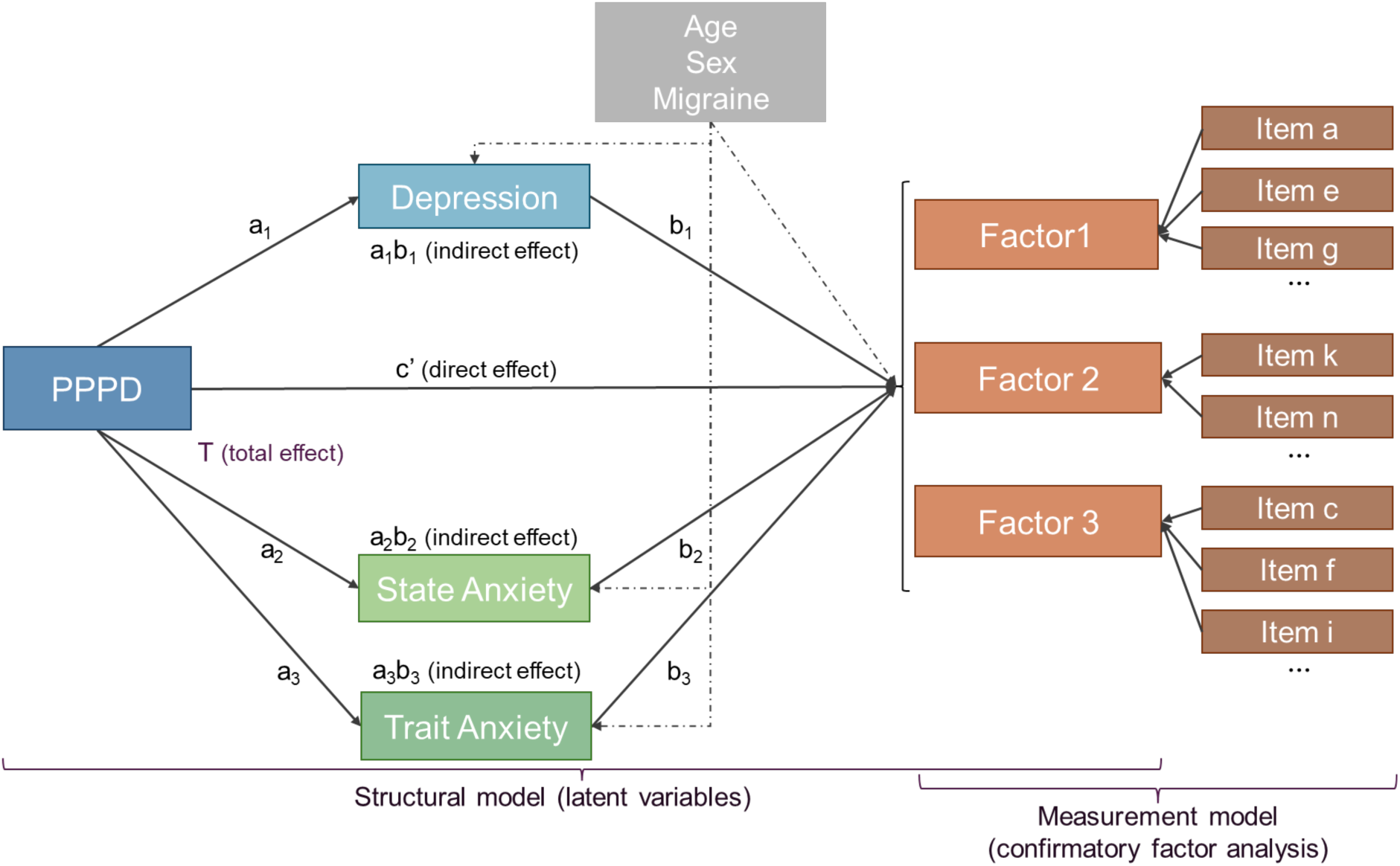
Structural equation model (SEM) testing the effect of persistent postural-perceptual dizziness (PPPD) on the three Cambridge Depersonalization Scale (CDS) factors. The model includes direct effects of PPPD (*vs* other otoneurological disorders) on each CDS factor, as well as indirect pathways mediated by depression, state anxiety, and trait anxiety. Age, sex, and migraine are included as covariates. The measurement model corresponds to the confirmatory factor analysis linking the three latent CDS factors to their respective items.

## Supplementary Results

**Supplementary Table 2.** Otoneurological diagnoses in participants with persistent postural-perceptual dizziness (PPPD) and other otoneurological disorders (OD).

| Form of vertigo/dizziness | Etiology | PPPD<br>*n* (%) | OD<br>*n* (%) |
| --- | --- | --- | --- |
| **Peripheral** | Benign paroxysmal positional vertigo (BPPV) | 8 (13.8%) | 49 (24.9%) |
|  | Menière's disease / Hydrops | 2 (3.5%) | 29 (14.7%) |
|  | Labyrinthitis | 3 (5.2%) | 10 (5.1%) |
|  | Perilymphatic fistula / Inner ear malformation | 1 (1.7%) | 12 (6.1%) |
|  | Acute unilateral vestibulopathy | 2 (3.5%) | 1 (0.5%) |
|  | Bilateral vestibulopathy | 1 (1.7%) | 13 (6.6%) |
|  | Vestibular paroxysmia | 2 (3.5%) | 4 (2.1%) |
|  | Vestibular schwannoma | 1 (1.7%) | 1 (0.5%) |
|  | Other unilateral vestibulopathy | 3 (5.2%) | 7 (3.5%) |
|  | Other unspecified vestibulopathy | 24 (41.3%) | 44 (22.3%) |
| **Central** | Vestibular migraine | 11 (18.9%) | 22 (11.2%) |
|  | Vestibular epilepsy | 0 (0%) | 3 (1.5%) |
| **Other** | Other | 0 (0%) | 2 (1.0%) |
| **All forms of vertigo/dizziness** |  | **54 (54%)** | **180 (100%)** |

The table lists past and/or current otoneurological comorbidities in PPPD patients and primary diagnoses in OD patients. Totals exceed the number of individuals because some patients had multiple co-occurring diagnoses.

**Supplementary Table 3.**
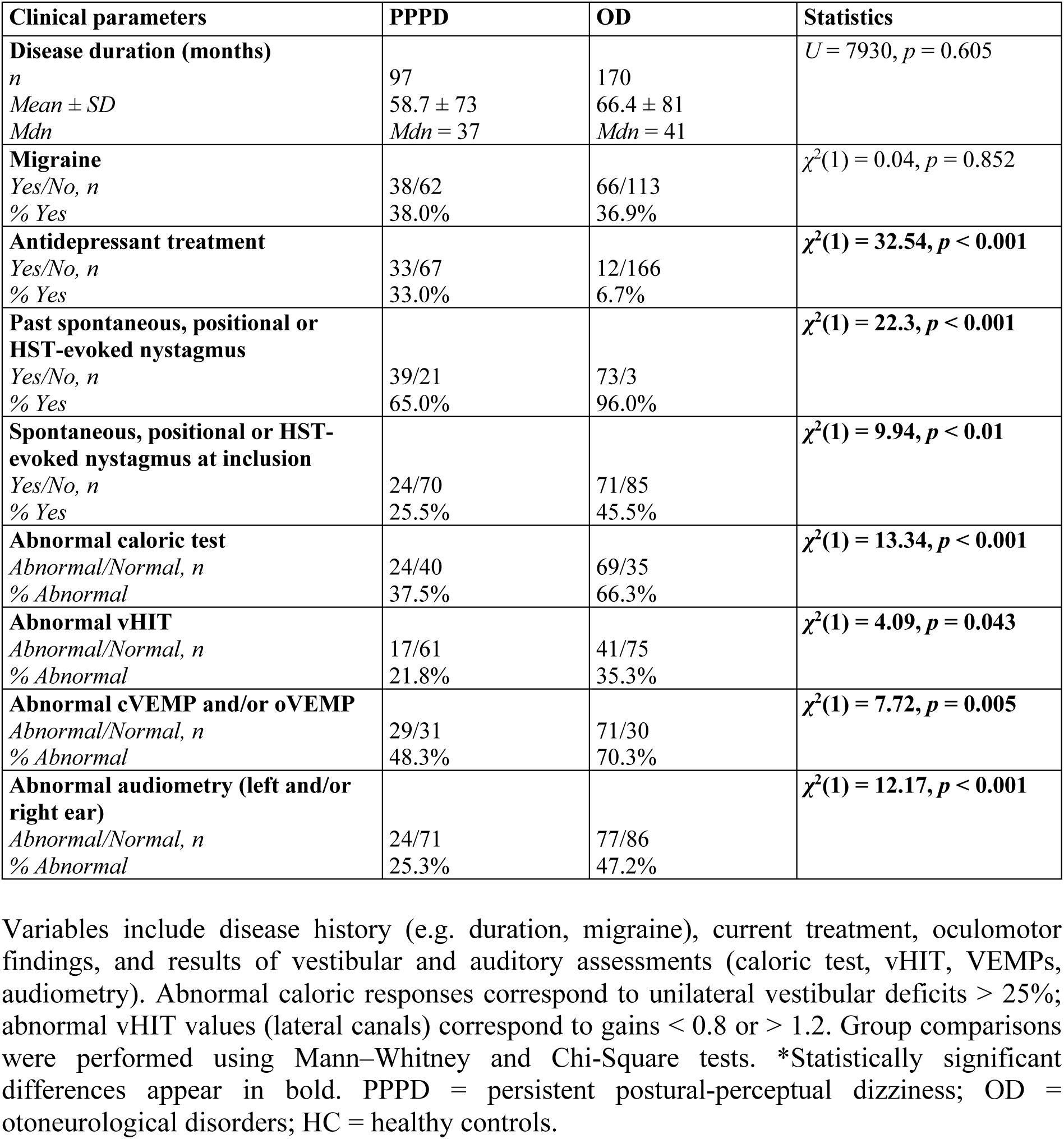
Descriptive statistics of clinical and vestibular function parameters.

| Clinical parameters | PPPD | OD | Statistics |
| --- | --- | --- | --- |
| **Disease duration (months)**<br>*n*<br>*Mean ± SD*<br>*Mdn* | 97<br>58.7 ± 73<br>*Mdn* = 37 | 170<br>66.4 ± 81<br>*Mdn* = 41 | $U = 7930, p = 0.605$ |
| **Migraine**<br>*Yes/No, n*<br>*% Yes* | 38/62<br>38.0% | 66/113<br>36.9% | $\chi^2(1) = 0.04, p = 0.852$ |
| **Antidepressant treatment**<br>*Yes/No, n*<br>*% Yes* | 33/67<br>33.0% | 12/166<br>6.7% | $\chi^2(1) = 32.54, p < 0.001$ |
| **Past spontaneous, positional or HST-evoked nystagmus**<br>*Yes/No, n*<br>*% Yes* | 39/21<br>65.0% | 73/3<br>96.0% | $\chi^2(1) = 22.3, p < 0.001$ |
| **Spontaneous, positional or HST-evoked nystagmus at inclusion**<br>*Yes/No, n*<br>*% Yes* | 24/70<br>25.5% | 71/85<br>45.5% | $\chi^2(1) = 9.94, p < 0.01$ |
| **Abnormal caloric test**<br>*Abnormal/Normal, n*<br>*% Abnormal* | 24/40<br>37.5% | 69/35<br>66.3% | $\chi^2(1) = 13.34, p < 0.001$ |
| **Abnormal vHIT**<br>*Abnormal/Normal, n*<br>*% Abnormal* | 17/61<br>21.8% | 41/75<br>35.3% | $\chi^2(1) = 4.09, p = 0.043$ |
| **Abnormal cVEMP and/or oVEMP**<br>*Abnormal/Normal, n*<br>*% Abnormal* | 29/31<br>48.3% | 71/30<br>70.3% | $\chi^2(1) = 7.72, p = 0.005$ |
| **Abnormal audiometry (left and/or right ear)**<br>*Abnormal/Normal, n*<br>*% Abnormal* | 24/71<br>25.3% | 77/86<br>47.2% | $\chi^2(1) = 12.17, p < 0.001$ |

Variables include disease history (e.g. duration, migraine), current treatment, oculomotor findings, and results of vestibular and auditory assessments (caloric test, vHIT, VEMPs, audiometry). Abnormal caloric responses correspond to unilateral vestibular deficits > 25%; abnormal vHIT values (lateral canals) correspond to gains < 0.8 or > 1.2. Group comparisons were performed using Mann–Whitney and Chi-Square tests. \*Statistically significant differences appear in bold. PPPD = persistent postural-perceptual dizziness; OD = otoneurological disorders; HC = healthy controls.

**Supplementary Table 4.** Descriptive statistics for the Cambridge Depersonalization Scale (CDS) total score and its factors.

| Subscale | Group | Median [IQR] | Mean ± SD | Min–Max |
| --- | --- | --- | --- | --- |
| **Total CDS score** | PPPD | 30 [13.8–60.5] | 42.6 ± 38.5 | 0–165 |
| | OD | 20 [7–37] | 26.7 ± 27.2 | 0–169 |
| | HC | 7 [2–17.5] | 12.5 ± 14.9 | 0–92 |
| **Frequency** | PPPD | 12 [6–22] | 16.6 ± 14.6 | 0–67 |
| | OD | 8 [3–14.2] | 10.5 ± 10.3 | 0–53 |
| | HC | 3 [1–7] | 5.3 ± 6.2 | 0–40 |
| **Duration** | PPPD | 17.5 [7.8–35.8] | 26.0 ± 24.5 | 0–98 |
| | OD | 11 [4–23.2] | 16.3 ± 17.7 | 0–120 |
| | HC | 4 [1–10] | 7.2 ± 9 | 0–52 |
| **Factor 1:**<br>**Bodily self disturbances** | PPPD | 1.8 [0.5–2.9] | 2.1 ± 2 | 0–7.8 |
| | OD | 0.8 [0–1.8] | 1.1 ± 1.4 | 0–8.6 |
| | HC | 0 [0–0.5] | 0.4 ± 0.7 | 0–3.6 |
| **Factor 2:**<br>**Cognitive and affective detachment** | PPPD | 0.6 [0.3–1.9] | 1.3 ± 1.5 | 0–6.1 |
| | OD | 0.7 [0.3–1.3] | 1.0 ± 1.2 | 0–8.3 |
| | HC | 0.6 [0.3–1] | 0.7 ± 0.7 | 0–4 |
| **Factor 3:**<br>**Numbing** | PPPD | 0 [0–2.1] | 1.3 ± 1.9 | 0–8 |
| | OD | 0 [0–1.3] | 1.0 ± 1.8 | 0–9 |
| | HC | 0 [0–0] | 0.3 ± 0.9 | 0–5.3 |

Subscale scores were normalized by the number of items included in each factor. Values are reported as median [interquartile range (IQR)], mean ± standard deviation (SD), and minimum–maximum (Min–Max). PPPD = persistent postural-perceptual dizziness; OD = otoneurological disorders; HC = healthy controls. For all measures, sample sizes are PPPD = 100, OD = 180, and HC = 175.

**Supplementary Table 5.**
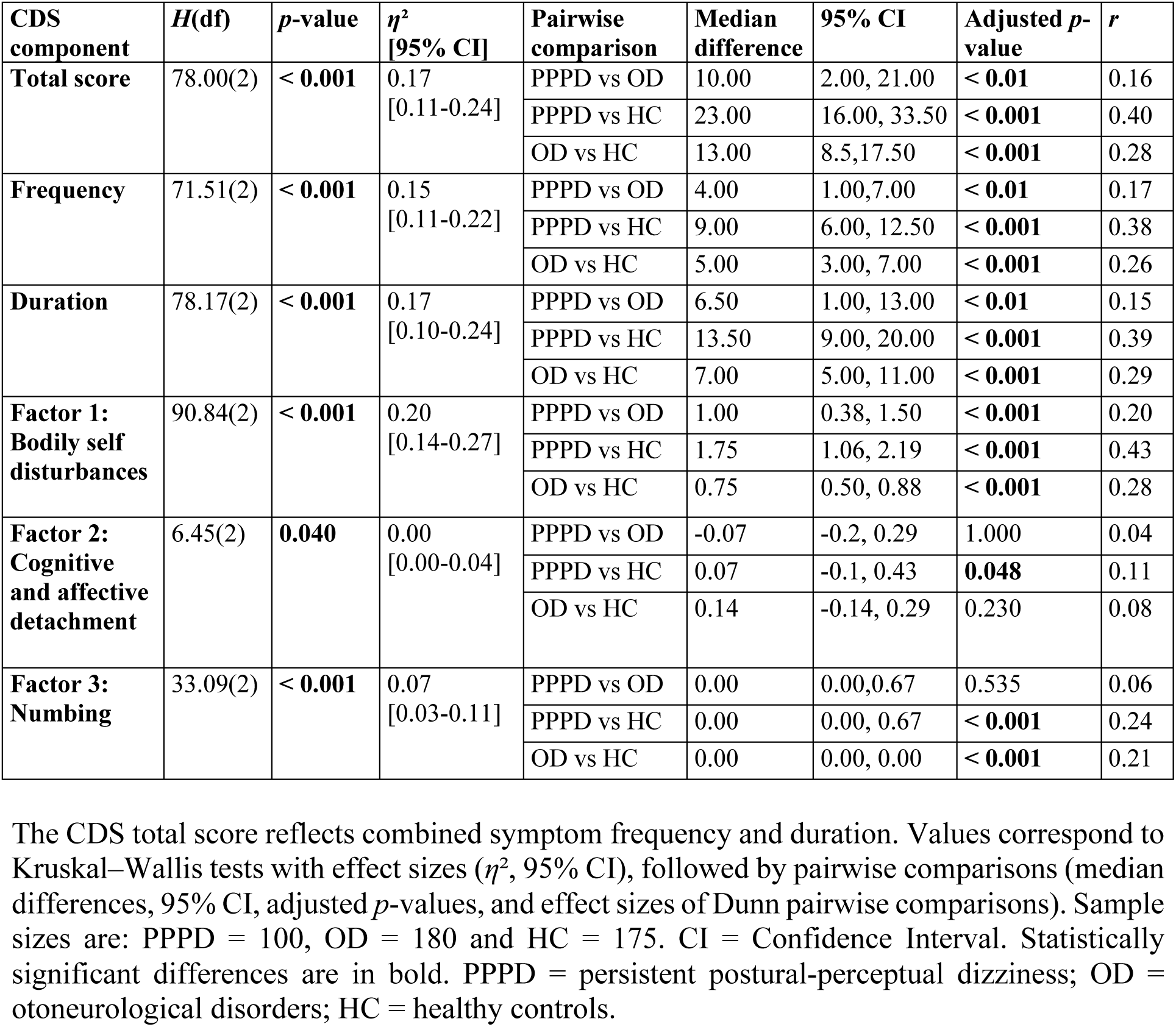
Intergroup comparisons of the Cambridge Depersonalization Scale (CDS) total score and its factors.

| CDS component | *H*(df) | *p*-value | $\eta^2$<br>[95% CI] | Pairwise comparison | Median difference | 95% CI | Adjusted *p*-value | *r* |
| --- | --- | --- | --- | --- | --- | --- | --- | --- |
| **Total score** | 78.00(2) | < **0.001** | 0.17<br>[0.11-0.24] | PPPD vs OD | 10.00 | 2.00, 21.00 | < **0.01** | 0.16 |
|  |  |  |  | PPPD vs HC | 23.00 | 16.00, 33.50 | < **0.001** | 0.40 |
|  |  |  |  | OD vs HC | 13.00 | 8.5, 17.50 | < **0.001** | 0.28 |
| **Frequency** | 71.51(2) | < **0.001** | 0.15<br>[0.11-0.22] | PPPD vs OD | 4.00 | 1.00, 7.00 | < **0.01** | 0.17 |
|  |  |  |  | PPPD vs HC | 9.00 | 6.00, 12.50 | < **0.001** | 0.38 |
|  |  |  |  | OD vs HC | 5.00 | 3.00, 7.00 | < **0.001** | 0.26 |
| **Duration** | 78.17(2) | < **0.001** | 0.17<br>[0.10-0.24] | PPPD vs OD | 6.50 | 1.00, 13.00 | < **0.01** | 0.15 |
|  |  |  |  | PPPD vs HC | 13.50 | 9.00, 20.00 | < **0.001** | 0.39 |
|  |  |  |  | OD vs HC | 7.00 | 5.00, 11.00 | < **0.001** | 0.29 |
| **Factor 1: Bodily self disturbances** | 90.84(2) | < **0.001** | 0.20<br>[0.14-0.27] | PPPD vs OD | 1.00 | 0.38, 1.50 | < **0.001** | 0.20 |
|  |  |  |  | PPPD vs HC | 1.75 | 1.06, 2.19 | < **0.001** | 0.43 |
|  |  |  |  | OD vs HC | 0.75 | 0.50, 0.88 | < **0.001** | 0.28 |
| **Factor 2: Cognitive and affective detachment** | 6.45(2) | **0.040** | 0.00<br>[0.00-0.04] | PPPD vs OD | -0.07 | -0.2, 0.29 | 1.000 | 0.04 |
|  |  |  |  | PPPD vs HC | 0.07 | -0.1, 0.43 | **0.048** | 0.11 |
|  |  |  |  | OD vs HC | 0.14 | -0.14, 0.29 | 0.230 | 0.08 |
| **Factor 3: Numbing** | 33.09(2) | < **0.001** | 0.07<br>[0.03-0.11] | PPPD vs OD | 0.00 | 0.00, 0.67 | 0.535 | 0.06 |
|  |  |  |  | PPPD vs HC | 0.00 | 0.00, 0.67 | < **0.001** | 0.24 |
|  |  |  |  | OD vs HC | 0.00 | 0.00, 0.00 | < **0.001** | 0.21 |

The CDS total score reflects combined symptom frequency and duration. Values correspond to Kruskal–Wallis tests with effect sizes ( $\eta^2$ , 95% CI), followed by pairwise comparisons (median differences, 95% CI, adjusted *p*-values, and effect sizes of Dunn pairwise comparisons). Sample sizes are: PPPD = 100, OD = 180 and HC = 175. CI = Confidence Interval. Statistically significant differences are in bold. PPPD = persistent postural-perceptual dizziness; OD = otoneurological disorders; HC = healthy controls.

**Supplementary Table 6.** Twenty-nine questionnaire items from the Cambridge Depersonalization in the order they were administered.

| CDS item | | *H*(df) | *p*-value | $\eta^2$ | Post-hoc pairwise comparison |
| --- | --- | --- | --- | --- | --- |
| Q1 | Out of the blue, I feel strange, as if I were not real or as if I were cut off from the world. | 98.66(2) | < **0.00172** | 0.214 | PPPD vs OD: MD = 1 [0; 3], *p* < **0.01**<br>PPPD vs HC: MD = 3 [2; 3], *p* < **0.001**<br>OD vs HC: MD = 2 [0; 2], *p* < **0.001** |
| Q2 | What I see looks ‘flat’ or ‘lifeless’, as if I were looking at a picture. | 26.01(2) | < **0.00172** | 0.053 | PPPD vs OD: MD = 0 [0; 0], *p* < **0.01**<br>PPPD vs HC: MD = 0 [0; 0], *p* < **0.001**<br>OD vs HC: MD = 0 [0; 0], *p* = 0.130 |
| Q3 | Parts of my body feel as if they didn’t belong to me. | 40.98(2) | < **0.00172** | 0.086 | PPPD vs OD: MD = 0 [0; 0], *p* < **0.05**<br>PPPD vs HC: MD = 0 [0; 0], *p* < **0.001**<br>OD vs HC: MD = 0 [0; 0], *p* < **0.001** |
| Q4 | I have found myself *not being frightened at all* in situations which normally I would find frightening or distressing. | 1.30(2) | 0.522 | -0.002 |  |
| Q5 | My favourite activities are no longer enjoyable. | 76.30(2) | < **0.00172** | 0.164 | PPPD vs OD: MD = 2 [1; 3], *p* < **0.01**<br>PPPD vs HC: MD = 4 [4; 5], *p* < **0.001**<br>OD vs HC: MD = 2 [2; 3], *p* < **0.001** |
| Q6 | Whilst doing something I have the feeling of being a ‘detached observer’ of myself. | 49.60(2) | < **0.00172** | 0.105 | PPPD vs OD: MD = 2 [0; 3], *p* < **0.001**<br>PPPD vs HC: MD = 2 [0; 3], *p* < **0.001**<br>OD vs HC: MD = 0 [0; 0], *p* < **0.01** |
| Q7 | The flavour of meals no longer gives me a feeling of pleasure or distaste. | 29.33(2) | < **0.00172** | 0.061 | PPPD vs OD: MD = 0 [0; 0], *p* = 0.846<br>PPPD vs HC: MD = 0 [0; 0], *p* < **0.001**<br>OD vs HC: MD = 0 [0; 0], *p* < **0.001** |
| Q8 | My body feels very light, as if it were floating on air. | 38.02(2) | < **0.00172** | 0.080 | PPPD vs OD: MD = 0 [0; 2], *p* < **0.01**<br>PPPD vs HC: MD = 0 [0; 2], *p* < **0.001**<br>OD vs HC: MD = 0 [0; 0], *p* < **0.001** |
| Q9 | When I weep or laugh, I do not seem *to feel* any emotions at all. | 13.52(2) | < **0.00172** | 0.026 | PPPD vs OD: MD = 0 [0; 0], *p* = 0.063<br>PPPD vs HC: MD = 0 [0; 0], *p* < **0.001**<br>OD vs HC: MD = 0 [0; 0], *p* = 0.310 |
| Q10 | I have the feeling of *not having any thoughts at all*, so that when I speak it feels as if my words were being uttered by an ‘automaton’. | 17.85(2) | < **0.00172** | 0.035 | PPPD vs OD: MD = 0 [0; 0], *p* < **0.05**<br>PPPD vs HC: MD = 0 [0; 0], *p* < **0.001**<br>OD vs HC: MD = 0 [0; 0], *p* = 0.355 |
| Q11 | Familiar voices (including my own) sound remote and unreal. | 22.98(2) | < **0.00172** | 0.046 | PPPD vs OD: MD = 0 [0; 0], *p* < **0.05**<br>PPPD vs HC: MD = 0 [0; 0], *p* < **0.01**<br>OD vs HC: MD = 0 [0; 0], *p* < **0.05** |
| Q12 | I have the feeling that my hands or my feet have become larger or smaller. | 15.30(2) | < **0.00172** | 0.029 | PPPD vs OD: MD = 0 [0; 0], *p* < **0.01**<br>PPPD vs HC: MD = 0 [0; 0], *p* < **0.01**<br>OD vs HC: MD = 0 [0; 0], *p* = 1.000 |
| Q13 | My surroundings feel detached or unreal, as if there were a veil between me and the outside world. | 38.88(2) | < **0.00172** | 0.082 | PPPD vs OD: MD = 1 [0; 3], *p* < **0.001**<br>PPPD vs HC: MD = 1 [0; 3], *p* < **0.001**<br>OD vs HC: MD = 0 [0; 0], *p* < **0.05** |
| Q14 | It seems as if things that I have recently done had taken place a long time ago. For example, anything which I have done this morning feels as if it were done weeks ago. | 2.10(2) | 0.350 | 0.000 |  |
| Q15 | Whilst fully awake I have ‘visions’ in which I can *see* myself outside, as if I were looking my image in a mirror. | 1.52(2) | 0.468 | -0.001 |  |
| Q16 | I feel detached from memories of things that have happened to me – as if I had not been involved in them. | 7.18(2) | 0.028 | 0.011 |  |

| CDS item | *H*(df) | *p*-value | $\eta^2$ | Post-hoc pairwise comparison |
| --- | --- | --- | --- | --- |
| Q17 When in a new situation, it feels as if I have been through it before. | 1.35(2) | 0.510 | -0.001 |  |
| Q18 Out of the blue, I find myself not feeling any affection towards my family and close friends. | 14.83(2) | < **0.00172** | 0.028 | PPPD vs OD: MD = 0 [0; 0], *p* = 0.430<br>PPPD vs HC: MD = 0 [0; 0], *p* < **0.001**<br>OD vs HC: MD = 0 [0; 0], *p* < **0.05** |
| Q19 Objects around me seem to look smaller or further away. | 19.38(2) | < **0.00172** | 0.038 | PPPD vs OD: MD = 0 [0; 0], *p* = 1.000<br>PPPD vs HC: MD = 0 [0; 0], *p* < **0.001**<br>OD vs HC: MD = 0 [0; 0], *p* < **0.01** |
| Q20 I cannot feel properly the objects that I touch with my hands for it feels *as if it were not me* who were touching it. | 11.02(2) | 0.004 | 0.020 |  |
| Q21 I do not seem able to picture things in my mind, for example, the face of a close friend or a familiar place. | 9.46(2) | 0.009 | 0.017 |  |
| Q22 When a part of my body hurts, I feel so detached from the pain that it feels as if it were ‘somebody else’s pain’. | 4.17(2) | 0.125 | 0.005 |  |
| Q23 I have the feeling of being outside my body. | 30.17(2) | < **0.00172** | 0.062 | PPPD vs OD: MD = 0 [0; 0], *p* < **0.001**<br>PPPD vs HC: MD = 0 [0; 0], *p* < **0.001**<br>OD vs HC: MD = 0 [0; 0], *p* = 0.423 |
| Q24 When I move it doesn’t feel as if I were in charge of the movements, so that I feel ‘automatic’ and mechanical as if I were a ‘robot’. | 53.28(2) | < **0.00172** | 0.113 | PPPD vs OD: MD = 2 [0; 3], *p* < **0.01**<br>PPPD vs HC: MD = 2 [0; 3], *p* < **0.001**<br>OD vs HC: MD = 0 [0; 0], *p* < **0.001** |
| Q25 The smell of things no longer gives me a feeling of pleasure or dislike. | 16.61(2) | < **0.00172** | 0.032 | PPPD vs OD: MD = 0 [0; 0], *p* = 0.139<br>PPPD vs HC: MD = 0 [0; 0], *p* < **0.001**<br>OD vs HC: MD = 0 [0; 0], *p* = 0.050 |
| Q26 I feel so detached from my thoughts that they seem to have a ‘life’ of their own. | 11.87(2) | 0.003 | 0.022 |  |
| Q27 I have to touch myself to make sure that I have a body or a real existence. | 11.55(2) | 0.003 | 0.021 |  |
| Q28 *I seem to have lost* some bodily sensations (e.g. of hunger and thirst) so that when I eat or drink, it feels an automatic routine. | 13.70(2) | < **0.00172** | 0.026 | PPPD vs OD: MD = 0 [0; 0], *p* = 0.386<br>PPPD vs HC: MD = 0 [0; 0], *p* < **0.01**<br>OD vs HC: MD = 0 [0; 0], *p* < **0.05** |
| Q29 Previously familiar places look unfamiliar, as if I had never seen them before. | 2.49(2) | 0.287 | 0.001 |  |

For each item, Kruskal–Wallis tests report *H*, degrees of freedom (df), *P*-value, and effect size ( $\eta^2$ ). Statistically significant group effects are in bold (adjusted statistical significance level: $\alpha_{\text{corr}} = 0.05/29 \approx 0.00172$ ). Post-hoc pairwise comparisons use Dunn tests with Bonferroni correction; median differences (MD) are presented with 95% confidence intervals (CI) and corrected *P*-values. Statistically significant between-group differences are shown in bold (adjusted *P*-value < 0.05). PPPD = persistent postural-perceptual dizziness; OD = otoneurological disorders; HC = healthy controls.

**Supplementary Figure 2.**
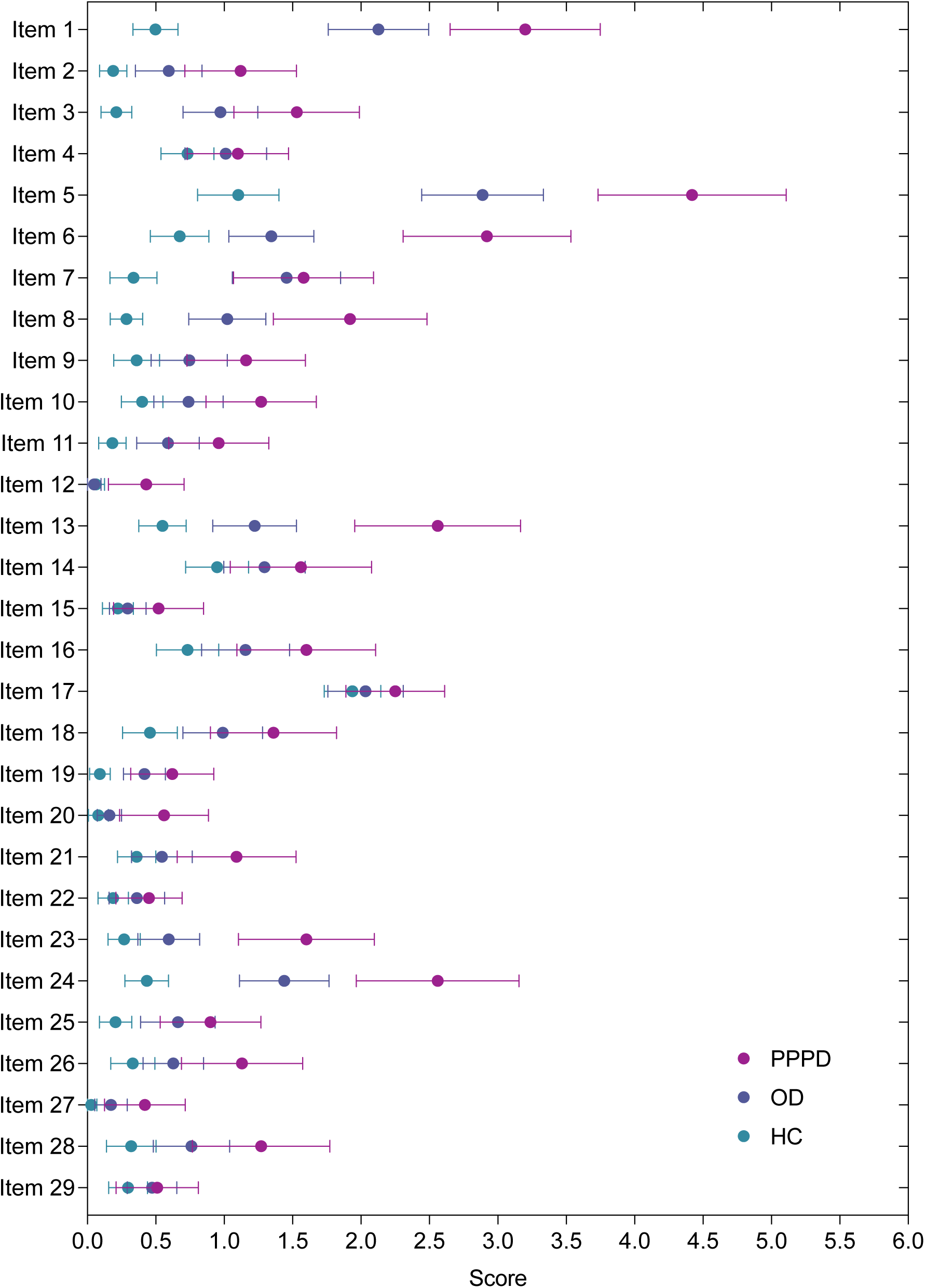
Comparison plot for the 29 Cambridge Depersonalization Scale item scores across groups. Dots represent group means and horizontal whiskers show the 95% confidence interval for each item. PPPD = persistent postural-perceptual dizziness; OD = otoneurological disorders; HC = healthy controls.

**Supplementary Table 7.**
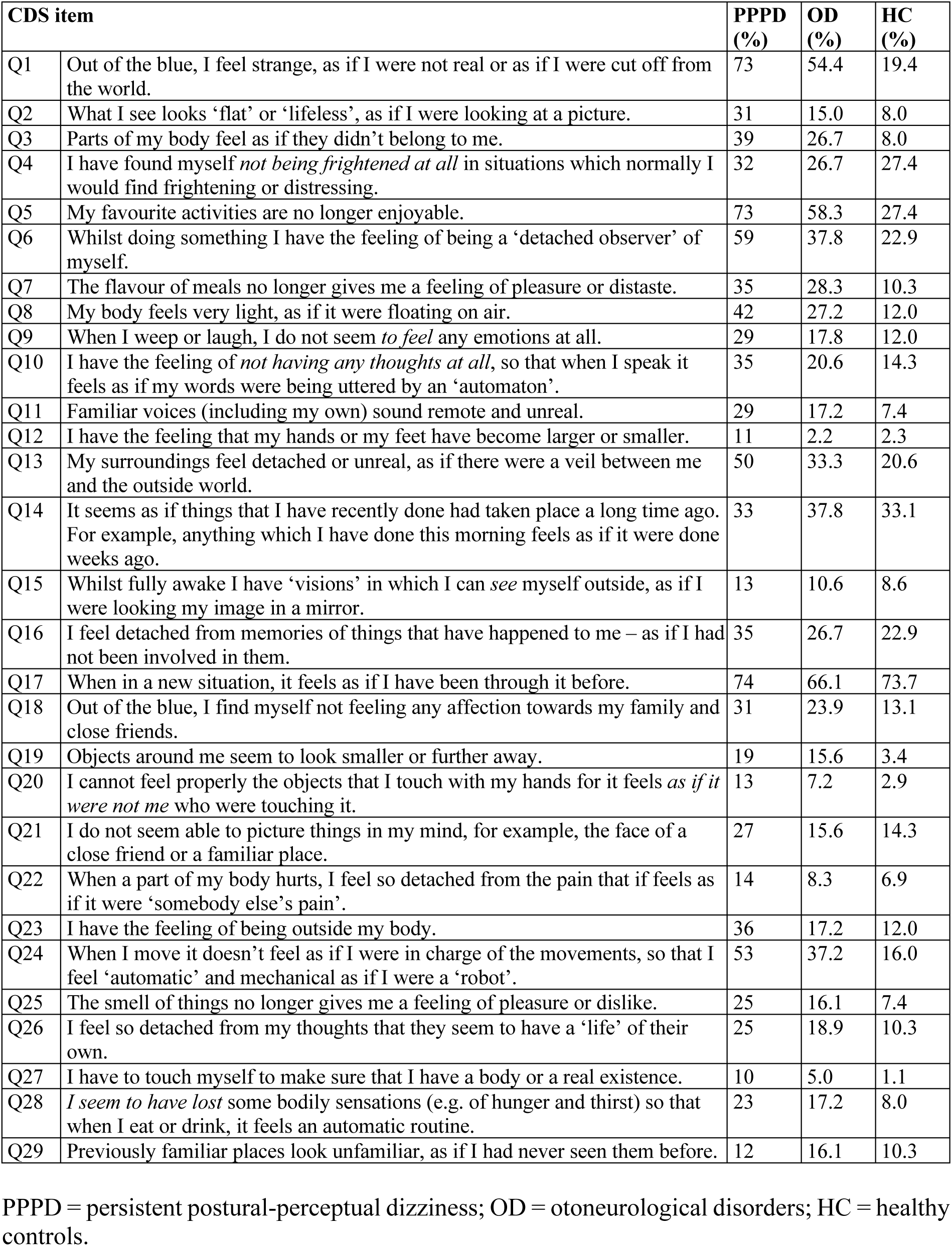
Proportion of individuals in each group reporting each of the 29 Cambridge Depersonalization Scale items.

**Supplementary Table 8.** Summary of the exploratory factor analysis and reliability results.

|  | **Factor 1** | **Factor 2** | **Factor 3** | **Factor 4** |
| --- | --- | --- | --- | --- |
| Number of items with factor loadings > 0.4 | 9 | 7 | 3 | 3 |
| Eigenvalues of factors | 9.14 | 1.37 | 1.05 | 0.78 |
| Percentage of variance | 16.7 | 9.9 | 6.8 | 6.2 |
| Cumulative Variance | 16.7 | 26.6 | 33.3 | 39.5 |
| Reliability analysis (Cronbach's $\alpha$) | 0.88 | 0.70 | 0.77 | 0.65 |

Exploratory factor analysis was justified by a significant Bartlett's test of sphericity ( $\chi^2_{(406)} = 5667.57, p < 0.001$ ) and an adequate Kaiser–Meyer–Olkin index (KMO = 0.91).

**Supplementary Table 9.** Exploratory factor analysis results.

| Item | Factor 1 | Factor 2 | Factor 3 | Factor 4 | Communality |
| --- | --- | --- | --- | --- | --- |
| Q1 | 0.819 | -0.109 | -0.117 | 0.031 | 0.505 |
| Q2 | 0.756 | -0.128 | -0.033 | -0.118 | 0.367 |
| Q3 | 0.779 | -0.090 | 0.006 | 0.011 | 0.537 |
| Q4 | 0.022 | 0.357 | 0.205 | -0.121 | 0.231 |
| Q5 | 0.415 | -0.043 | 0.148 | 0.161 | 0.347 |
| Q6 | 0.674 | 0.054 | 0.023 | 0.085 | 0.602 |
| Q7 | -0.130 | -0.067 | 0.846 | 0.195 | 0.645 |
| Q8 | 0.358 | -0.101 | 0.154 | 0.215 | 0.294 |
| Q9 | 0.310 | 0.192 | 0.357 | -0.211 | 0.441 |
| Q10 | 0.517 | 0.276 | 0.122 | -0.180 | 0.530 |
| Q11 | 0.349 | 0.165 | 0.146 | -0.092 | 0.286 |
| Q12 | 0.101 | -0.029 | 0.057 | 0.347 | 0.178 |
| Q13 | 0.787 | 0.008 | 0.041 | 0.044 | 0.710 |
| Q14 | -0.057 | 0.622 | -0.054 | 0.223 | 0.465 |
| Q15 | -0.034 | 0.204 | 0.006 | 0.529 | 0.393 |
| Q16 | 0.074 | 0.635 | -0.042 | 0.037 | 0.462 |
| Q17 | -0.017 | 0.539 | -0.022 | -0.029 | 0.253 |
| Q18 | 0.021 | 0.543 | 0.076 | 0.030 | 0.384 |
| Q19 | -0.069 | -0.033 | 0.092 | 0.706 | 0.466 |
| Q20 | 0.077 | 0.021 | -0.009 | 0.695 | 0.560 |
| Q21 | 0.263 | 0.290 | -0.116 | -0.014 | 0.184 |
| Q22 | -0.199 | 0.482 | 0.150 | 0.114 | 0.266 |
| Q23 | 0.621 | 0.055 | -0.076 | 0.220 | 0.581 |
| Q24 | 0.644 | -0.069 | 0.082 | 0.164 | 0.568 |
| Q25 | -0.043 | 0.031 | 0.770 | -0.014 | 0.577 |
| Q26 | 0.238 | 0.453 | 0.153 | -0.035 | 0.528 |
| Q27 | 0.298 | 0.167 | -0.197 | 0.174 | 0.207 |
| Q28 | 0.102 | 0.028 | 0.493 | 0.107 | 0.392 |
| Q29 | -0.160 | 0.745 | -0.071 | 0.016 | 0.387 |

For each item of the Cambridge Depersonalization Scale, the table presents loadings on the four extracted factors and the corresponding communalities. The cumulative variance explained by the factors is shown in the last column.

**Supplementary Table 10.** Correlations between the extracted factors.

| **Factor** | **Factor 1** | **Factor 2** | **Factor 3** | **Factor 4** |
| --- | --- | --- | --- | --- |
| **Factor 1** | *1.00* | 0.66 | 0.59 | 0.56 |
| **Factor 2** | 0.66 | *1.00* | 0.60 | 0.45 |
| **Factor 3** | 0.59 | 0.60 | *1.00* | 0.29 |
| **Factor 4** | 0.56 | 0.45 | 0.29 | *1.00* |

The table reports between-factor correlations for the four factors identified in the exploratory factor analysis. Item 5 was removed to maintain theoretical coherence within Factor 1. Factor 4 was excluded from subsequent analyses because it violated configural invariance across groups. The final confirmatory factor analysis was conducted on this adjusted model.

**Supplementary Table 11.** Confirmatory factor analysis fit statistics across models.

| **Model** | **$\chi^2$ (df)**<br>**scaled** | ***p*-value**<br>**scaled** | **CFI**<br>**robust** | **TLI**<br>**robust** | **RMSEA**<br>**robust**<br>**[95%CI]** | **SRMR** | **AIC** | **BIC** |
| --- | --- | --- | --- | --- | --- | --- | --- | --- |
| **Global** | 224.390<br>(132) | < 0.001 | 0.946 | 0.937 | 0.055<br>[0.042-0.067] | 0.055 | 29633.912 | 29794.603 |
| **Configural**<br>**invariance** | 664.851<br>(396) | <0 .001 | 0.867 | 0.846 | 0.081<br>[0.070-0.092] | 0.076 | 28194.283 | 28898.854 |
| **Metric**<br>**invariance** | 664.857<br>(426) | < 0.001 | 0.873 | 0.864 | 0.076<br>[0.065-0.087] | 0.091 | 28201.040 | 28782.002 |
| **Scalar**<br>**invariance** | 732.670<br>(456) | < 0.001 | 0.857 | 0.856 | 0.078<br>[0.068-0.089] | 0.097 | 28222.928 | 28680.281 |
| **Partial**<br>**scalar**<br>**invariance** | 695.087<br>(450) | < 0.001 | 0.873 | 0.870 | 0.074<br>[0.063-0.085] | 0.092 | 28180.756 | 28662.831 |

The table provides robust fit indices and information criteria for the global model and for successive invariance models. Partial scalar invariance was obtained by releasing intercepts of items 1, 10, and 23 according to modification indices $\geq 8$ . Robust fit indices were computed using a robust maximum likelihood estimator (MLR) to account for non-normality in the data. CFI = Comparative Fit Index; TLI = Tucker–Lewis Index; RMSEA = Root Mean Square Error of Approximation; CI = Confidence Interval; SRMR = Standardized Root Mean Square Residual; AIC = Akaike Information Criterion; BIC = Bayesian Information Criterion.

**Supplementary Table 12.**
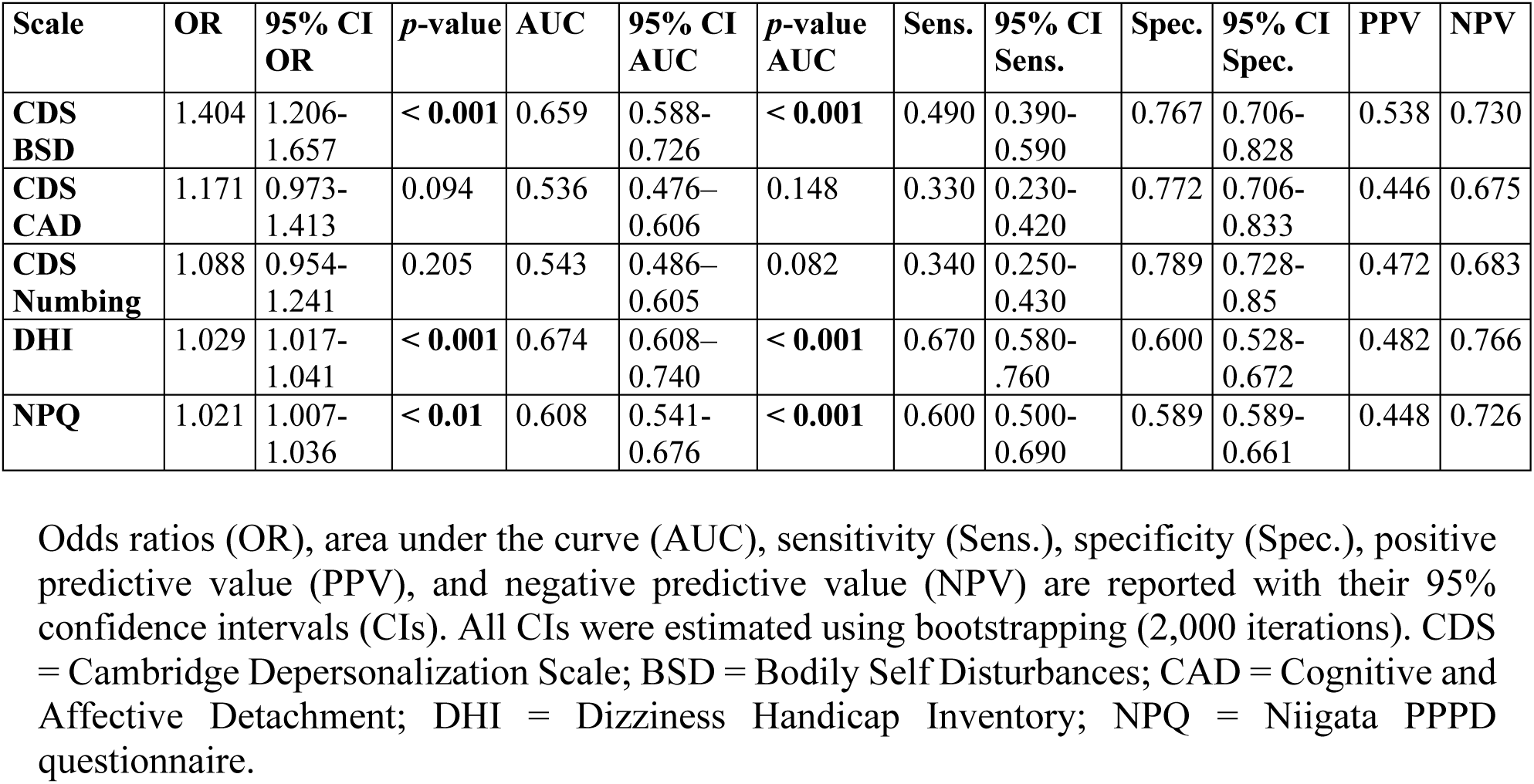
Receiver operating characteristic (ROC) curve analyses.

**Supplementary Table 13.** Pairwise comparisons of ROC curves between scales.

| Variable 1 | Variable 2 | AUC 1 | AUC 2 | AUC difference | 95% CI AUC difference | *p*-value |
| --- | --- | --- | --- | --- | --- | --- |
| CDS BSD | CDS CAD | 0.659 | 0.536 | 0.123 | 0.051 – 0.19 | < **0.01** |
| CDS BSD | CDS Numbing | 0.659 | 0.543 | 0.115 | 0.045 – 0.185 | < **0.001** |
| CDS CAD | CDS Numbing | 0.536 | 0.543 | -0.007 | -0.07 – 0.06 | 0.830 |
| CDS BSD | DHI | 0.659 | 0.674 | -0.016 | -0.084 – 0.052 | 0.650 |
| CDS BSD | NPQ | 0.659 | 0.608 | 0.049 | -0.025 – 0.12 | 0.177 |
| CDS CAD | DHI | 0.536 | 0.674 | -0.137 | -0.22 – -0.055 | < **0.01** |
| CDS CAD | NPQ | 0.536 | 0.608 | -0.072 | -0.157 – 0.013 | 0.093 |
| CDS Numbing | DHI | 0.543 | 0.674 | -0.131 | -0.206 – -0.058 | < **0.01** |
| CDS Numbing | NPQ | 0.543 | 0.608 | -0.066 | -0.145 – 0.01 | 0.087 |
| DHI | NPQ | 0.674 | 0.608 | 0.065 | 0.011 – 0.119 | < **0.05** |

Differences between areas under the curves (AUC) are presented with their 95% confidence intervals (CIs), together with *P*-values from bootstrap comparison of ROC curves. CDS = Cambridge Depersonalization Scale; BSD = Bodily Self Disturbances; CAD = Cognitive and Affective Detachment; DHI = Dizziness Handicap Inventory; NPQ = Niigata PPPD questionnaire.

**Supplementary Table 14.** Logistic regression across quantiles for CDS factors, NPQ and DHI.

| Scale | Quantile | *n*<br>total | *n*<br>PPPD | PPPD<br>proportion | 95% CI PPPD | OR | 95% CI OR | *p*-value |
| --- | --- | --- | --- | --- | --- | --- | --- | --- |
| CDS BSD | Quartile 1 | 70 | 15 | 0.214 | 0.129 – 0.332 | *Reference* |  |  |
|  | Quartile 2 | 70 | 20 | 0.286 | 0.187 – 0.408 | 1.467 | 0.681 – 3.213 | 0.330 |
|  | Quartile 3 | 70 | 27 | 0.386 | 0.274 – 0.510 | 2.302 | 1.103 – 4.948 | < **0.05** |
|  | Quartile 4 | 70 | 38 | 0.543 | 0.420 – 0.661 | 4.354 | 2.114 – 9.336 | < **0.001** |
|  | Bottom 10% | 28 | 5 | 0.179 | 0.068 – 0.376 | *Reference* |  |  |
|  | Top 10% | 28 | 19 | 0.679 | 0.476 – 0.834 | 9.711 | 2.959 – 37.157 | < **0.001** |
| CDS CAD | Quartile 1 | 70 | 23 | 0.329 | 0.224 – 0.452 | *Reference* |  |  |
|  | Quartile 2 | 70 | 27 | 0.386 | 0.274 – 0.510 | 1.283 | 0.642 – 2.580 | 0.481 |
|  | Quartile 3 | 70 | 19 | 0.271 | 0.175 – 0.393 | 0.761 | 0.366 – 1.570 | 0.461 |
|  | Quartile 4 | 70 | 31 | 0.443 | 0.326 – 0.566 | 1.624 | 0.821 – 3.251 | 0.166 |
|  | Bottom 10% | 28 | 5 | 0.179 | 0.068 – 0.376 | *Reference* |  |  |
|  | Top 10% | 28 | 15 | 0.536 | 0.342 – 0.720 | 5.308 | 1.650 – 19.541 | < **0.001** |
| CDS<br>Numbing | Quartile 1 | 70 | 16 | 0.229 | 0.140 – 0.347 | *Reference* |  |  |
|  | Quartile 2 | 70 | 20 | 0.286 | 0.187 – 0.408 | 1.350 | 0.632 – 2.922 | 0.440 |
|  | Quartile 3 | 70 | 31 | 0.443 | 0.326 – 0.566 | 2.683 | 1.307 – 5.67 | < **0.01** |
|  | Quartile 4 | 70 | 33 | 0.471 | 0.352 – 0.594 | 3.010 | 1.471 – 6.359 | < **0.01** |
|  | Bottom 10% | 28 | 10 | 0.357 | 0.193 – 0.559 | *Reference* |  |  |
|  | Top 10% | 28 | 13 | 0.464 | 0.280 – 0.658 | 1.560 | 0.537 – 4.645 | 0.416 |
| DHI | Quartile 1 | 70 | 9 | 0.129 | 0.064 – 0.235 | *Reference* |  |  |
|  | Quartile 2 | 70 | 24 | 0.343 | 0.236 – 0.467 | 3.536 | 1.546 – 8.71 | < **0.01** |
|  | Quartile 3 | 70 | 30 | 0.429 | 0.313 – 0.552 | 5.083 | 2.258 – 12.418 | < **0.001** |
|  | Quartile 4 | 70 | 37 | 0.529 | 0.406 – 0.648 | 7.599 | 3.393 – 18.56 | < **0.001** |
|  | Bottom 10% | 28 | 3 | 0.107 | 0.030 – 0.294 | *Reference* |  |  |
|  | Top 10% | 28 | 15 | 0.536 | 0.342 – 0.720 | 9.616 | 2.613 – 47.245 | < **0.01** |
| NPQ | Quartile 1 | 70 | 16 | 0.229 | 0.140 – 0.347 | *Reference* |  |  |
|  | Quartile 2 | 70 | 22 | 0.314 | 0.211 – 0.438 | 1.547 | 0.733 – 3.323 | 0.256 |
|  | Quartile 3 | 70 | 30 | 0.429 | 0.313 – 0.552 | 2.531 | 1.232 – 5.354 | < **0.05** |
|  | Quartile 4 | 70 | 32 | 0.457 | 0.339 – 0.580 | 2.842 | 1.387 – 6.005 | < **0.01** |
|  | Bottom 10% | 28 | 5 | 0.179 | 0.068 – 0.376 | *Reference* |  |  |
|  | Top 10% | 28 | 15 | 0.536 | 0.342 – 0.720 | 5.308 | 1.650 – 19.541 | < **0.01** |

For each scale, proportions of individuals with PPPD are reported by quartile and by the bottom and top deciles, together with their 95% confidence intervals (CIs). Odds ratios (ORs) compare each quantile with the corresponding reference category (Quartile 1 or Bottom 10%). CDS = Cambridge Depersonalization Scale; BSD = Bodily Self Disturbances; CAD = Cognitive and Affective Detachment; DHI = Dizziness Handicap Inventory; NPQ = Niigata PPPD questionnaire.

**Supplementary Table 15.**
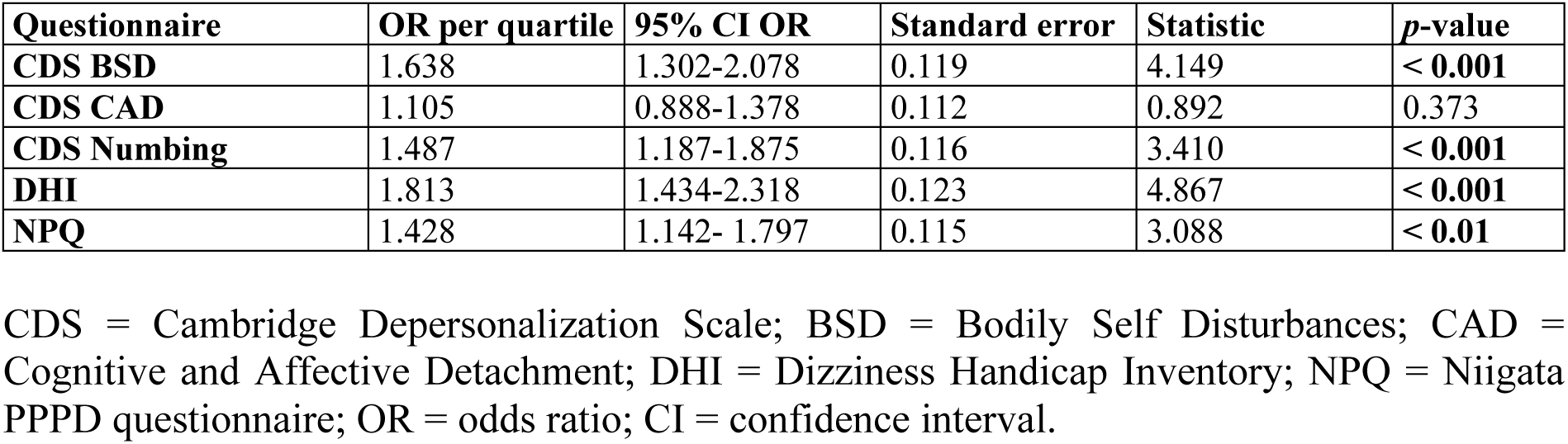
Linear trend test across quartiles.

**Supplementary Table 16.**
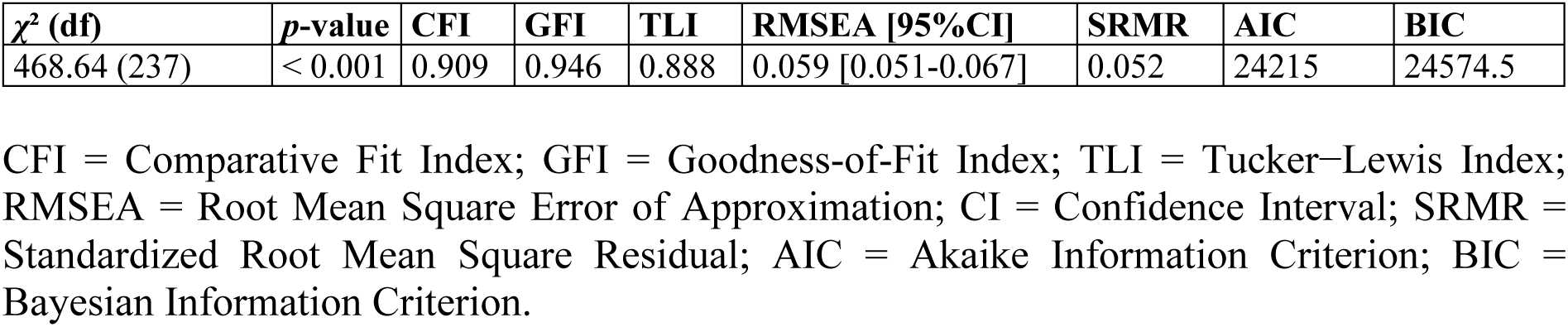
Results from structural equation modelling: fit indices.

| $\chi^2$ (df) | *p*-value | CFI | GFI | TLI | RMSEA [95%CI] | SRMR | AIC | BIC |
| --- | --- | --- | --- | --- | --- | --- | --- | --- |
| 468.64 (237) | < 0.001 | 0.909 | 0.946 | 0.888 | 0.059 [0.051-0.067] | 0.052 | 24215 | 24574.5 |

CFI = Comparative Fit Index; GFI = Goodness-of-Fit Index; TLI = Tucker–Lewis Index; RMSEA = Root Mean Square Error of Approximation; CI = Confidence Interval; SRMR = Standardized Root Mean Square Residual; AIC = Akaike Information Criterion; BIC = Bayesian Information Criterion.

**Supplementary Table 17.** Structural equation model: effect of PPPD, depression, state and trait anxiety on the different CDS factors for the whole clinical sample, as well as the effect of the covariates (migraine, age and sex) on the three CDS factors, depression, and anxiety.

| Factor | Predictor/Covariate | Std. Estimate (std.lv) | SE | z-value | p-value | 95% CI |
| --- | --- | --- | --- | --- | --- | --- |
| **Bodily self disturbances** | Depression | 0.080 | 0.019 | 4.223 | < **0.001** | 0.043, 0.117 |
|  | State Anxiety | -0.001 | 0.019 | -0.051 | 0.959 | -0.037, 0.036 |
|  | Trait Anxiety | 0.020 | 0.008 | 2.427 | < **0.05** | 0.004, 0.035 |
|  | PPPD | 0.319 | 0.131 | 2.438 | < **0.01** | 0.062, 0.575 |
|  | Age | -0.011 | 0.004 | -2.665 | < **0.01** | -0.018, -0.003 |
|  | Sex | 0.064 | 0.118 | 0.539 | 0.590 | -0.167, 0.294 |
|  | Migraine | 0.142 | 0.117 | 1.215 | 0.224 | -0.087, 0.371 |
| **Cognitive and affective detachment** | Depression | 0.084 | 0.021 | 4.075 | < **0.001** | 0.043, 0.124 |
|  | State Anxiety | -0.027 | 0.021 | -1.317 | 0.188 | -0.068, 0.013 |
|  | Trait Anxiety | 0.023 | 0.009 | 2.489 | < **0.05** | 0.005, 0.042 |
|  | PPPD | -0.044 | 0.130 | -0.341 | 0.733 | -0.300, 0.211 |
|  | Age | -0.008 | 0.005 | -1.491 | 0.136 | -0.019, 0.003 |
|  | Sex | 0.002 | 0.133 | 0.018 | 0.985 | -0.258, 0.263 |
|  | Migraine | 0.181 | 0.129 | 1.410 | 0.158 | -0.071, 0.433 |
| **Numbing** | Depression | 0.113 | 0.023 | 4.828 | < **0.001** | 0.067, 0.159 |
|  | State Anxiety | 0.027 | 0.023 | 1.165 | 0.244 | -0.019, 0.073 |
|  | Trait Anxiety | -0.009 | 0.010 | -0.959 | 0.337 | -0.029, 0.010 |
|  | PPPD | -0.100 | 0.136 | -0.732 | 0.464 | -0.367, 0.167 |
|  | Age | -0.002 | 0.005 | -0.472 | 0.637 | -0.012, 0.007 |
|  | Sex | 0.034 | 0.139 | 0.244 | 0.807 | -0.238, 0.306 |
|  | Migraine | 0.250 | 0.137 | 1.818 | 0.069 | -0.019, 0.519 |
| **Depression** | PPPD | 1.802 | 0.498 | 3.620 | < **0.001** | 0.826, 2.778 |
|  | Age | -0.045 | 0.018 | -2.513 | < **0.05** | -0.080, -0.010 |
|  | Sex | -0.871 | 0.523 | -1.667 | 0.096 | -1.896, 0.153 |
|  | Migraine | 1.104 | 0.489 | 2.257 | < **0.05** | 0.145, 2.062 |
| **State Anxiety** | PPPD | 0.784 | 0.517 | 1.516 | 0.129 | -0.229, 1.797 |
|  | Age | -0.078 | 0.020 | -3.904 | < **0.001** | -0.117, -0.039 |
|  | Sex | 1.266 | 0.522 | 2.425 | < **0.05** | 0.243, 2.289 |
|  | Migraine | 1.355 | 0.497 | 2.726 | < **0.001** | 0.381, 2.330 |
| **Trait Anxiety** | PPPD | 3.312 | 1.390 | 2.382 | < **0.05** | 0.587, 6.037 |
|  | Age | -0.191 | 0.048 | -3.972 | < **0.001** | -0.285, -0.097 |
|  | Sex | 0.543 | 1.534 | 0.354 | 0.723 | -2.463, 3.550 |
|  | Migraine | 1.291 | 1.367 | 0.945 | 0.345 | -1.388, 3.970 |

PPPD = PPPD vs. OD; Std. Estimate (std.lv) = standardized coefficient estimate for latent variables; SE = standard error; CI = confidence interval. Reference for Sex = Male.

**Supplementary Table 18.** Structural equation model: indirect and total effects of PPPD on the three CDS factors mediated by depression, state anxiety, and trait anxiety in the whole clinical sample.

| Predictor | Mediator | Outcome | Effect type | Std. Estimate (std.lv) | SE | z | p-value | 95% CI |
| --- | --- | --- | --- | --- | --- | --- | --- | --- |
| PPPD | Depression | Bodily self disturbances | Indirect | 0.144 | 0.053 | 2.715 | < **0.01** | 0.040, 0.249 |
|  | State Anxiety |  | Indirect | -0.001 | 0.015 | -0.051 | 0.959 | -0.029, 0.028 |
|  | Trait Anxiety |  | Indirect | 0.065 | 0.039 | 1.651 | 0.099 | -0.012, 0.142 |
|  |  |  | Total | 0.527 | 0.136 | 3.876 | < **0.001** | 0.261, 0.794 |
| PPPD | Depression | Cognitive and affective detachment | Indirect | 0.151 | 0.059 | 2.560 | < **0.01** | 0.035, 0.266 |
|  | State Anxiety |  | Indirect | -0.022 | 0.022 | -0.996 | 0.319 | -0.064, 0.021 |
|  | Trait Anxiety |  | Indirect | 0.077 | 0.043 | 1.793 | 0.073 | -0.007, 0.161 |
|  |  |  | Total | 0.162 | 0.155 | 1.045 | 0.296 | -0.142, 0.465 |
| PPPD | Depression | Numbing | Indirect | 0.204 | 0.069 | 2.947 | < **0.01** | 0.068, 0.339 |
|  | State Anxiety |  | Indirect | 0.021 | 0.023 | 0.928 | 0.353 | -0.024, 0.067 |
|  | Trait Anxiety |  | Indirect | -0.031 | 0.034 | -0.917 | 0.359 | -0.098, 0.036 |
|  |  |  | Total | 0.094 | 0.150 | 0.628 | 0.530 | -0.200, 0.388 |

PPPD = PPPD vs. OD; Std. Estimate (std.lv) = standardized factor loading; SE = standard error; CI = confidence interval.

**Supplementary Table 19.** Structural equation model: measurement model showing item loadings for the three CDS factors.

| Factor | Item | Std. Estimate (std.lv) | SE | z | p-value | 95% CI |
| --- | --- | --- | --- | --- | --- | --- |
| **Bodily self disturbances** | Q1 | 1.692 | 0.165 | 10.262 | < **0.001** | 1.369, 2.016 |
|  | Q2 | 0.963 | 0.197 | 4.886 | < **0.001** | 0.577, 1.350 |
|  | Q3 | 1.377 | 0.186 | 7.398 | < **0.001** | 1.012, 1.742 |
|  | Q6 | 2.061 | 0.159 | 12.942 | < **0.001** | 1.749, 2.374 |
|  | Q10 | 1.110 | 0.197 | 5.623 | < **0.001** | 0.723, 1.497 |
|  | Q13 | 2.133 | 0.176 | 12.125 | < **0.001** | 1.788, 2.478 |
|  | Q23 | 1.495 | 0.202 | 7.391 | < **0.001** | 1.099, 1.892 |
|  | Q24 | 1.864 | 0.192 | 9.707 | < **0.001** | 1.488, 2.241 |
| **Cognitive and affective detachment** | Q14 | 1.480 | 0.200 | 7.414 | < **0.001** | 1.089, 1.871 |
|  | Q16 | 1.680 | 0.205 | 8.190 | < **0.001** | 1.278, 2.083 |
|  | Q17 | 0.845 | 0.178 | 4.737 | < **0.001** | 0.495, 1.194 |
|  | Q18 | 1.278 | 0.173 | 7.404 | < **0.001** | 0.939, 1.616 |
|  | Q22 | 0.647 | 0.196 | 3.306 | < **0.001** | 0.264, 1.031 |
|  | Q26 | 1.252 | 0.208 | 6.024 | < **0.001** | 0.844, 1.659 |
|  | Q29 | 0.838 | 0.176 | 4.761 | < **0.001** | 0.493, 1.183 |
| **Numbing** | Q7 | 2.020 | 0.217 | 9.320 | < **0.001** | 1.596, 2.445 |
|  | Q25 | 1.287 | 0.222 | 5.800 | < **0.001** | 0.852, 1.722 |
|  | Q28 | 1.355 | 0.237 | 5.712 | < **0.001** | 0.890, 1.820 |

PPPD = PPPD vs. OD; Std. Estimate (std.lv) = standardized factor loading; SE = standard error; CI = confidence interval.

